# Human Kidney Alternative Splicing Information Illuminates Cardiovascular-Kidney-Metabolic Syndrome Risk

**DOI:** 10.1101/2025.10.02.25337157

**Authors:** Samer Mohandes, Daigoro Hirohama, Eunji Ha, Bernhard Dumoulin, Siyu Pan, Andrea Bergeson, Amin Abedini, Thao Nguyen, Fiona Elizabeth McAllister, TRIDENT Consortium, Katalin Susztak

## Abstract

Alternative splicing generates multiple transcripts from a single gene, contributing to functional protein diversity. However, short-read RNA sequencing often aggregates isoforms, limiting transcript-level resolution. By analyzing splicing variation in 404 human kidney cortical samples across a spectrum of disease severity, we define the splicing landscape of the human kidney in health and disease. To pinpoint likely disease-causing events, we performed splicing and expression quantitative trait locus (sQTL and eQTL) analyses and identified 1,948 genes with significant sQTLs. Integration with genome-wide association study (GWAS) data for kidney function and cardiovascular-kidney-metabolic (CKM) syndrome traits revealed 30 splice variants with evidence of shared genetic regulation via Bayesian colocalization and summary-based Mendelian randomization. Six genes—including MANBA, CHMP1A, NT5DC2, GSTA1, GSTA2, and MGMT—demonstrated concordant sQTL, eQTL, and protein QTL (pQTL) signals, implicating transcript-level regulation as a key mediator of kidney function–associated variation. Similarly, multi-layer QTL evidence implicated a *CLCNK* splice variant in blood pressure regulation. To resolve full transcript structures, we applied long-read RNA sequencing, identifying novel isoforms and validating short-read quantifications. These analyses nominate specific isoforms, including those of *MANBA* and *GSTA2*, as candidate mediators of kidney disease risk. Our findings highlight transcript-specific genetic regulation in the human kidney and underscore the value of isoform-resolved analysis for understanding complex trait biology.

## Introduction

Chronic kidney disease (CKD) affects approximately one in ten adults worldwide and is a major contributor to years of life lost^1,2^. Importantly, CKD rarely develops in isolation; impaired kidney function often evolves alongside obesity, diabetes, dyslipidemia, and hypertension—conditions now grouped under the umbrella of cardiovascular-kidney-metabolic (CKM) syndrome^3^. A deeper understanding of the molecular pathways that govern CKM traits has the potential to translate inherited risk into metabolic dysfunction, nephron loss or heart disease paving the development of curative interventions.

Although CKM syndrome is still incompletely understood, genetic studies focused on individual cardiac, kidney, and metabolic traits have built a strong foundation for integrative analyses across disease domains. Twin and family studies estimate that nearly half of the variance in individual’s estimated glomerular filtration rate (eGFR) is heritable^4^, motivating extensive genome-wide association studies (GWAS) for kidney traits. Although GWAS have identified over a thousand loci linked to estimated GFR and CKD^5^, more than 95 % of genetic variants reside in non-coding regions making it difficult to identify the causal genes^6^. Expression-quantitative trait locus (eQTL) mapping has helped bridge some of this gap by linking non-coding variants to changes in kidney gene expression; however, fewer than half of renal GWAS associations can be explained solely by total mRNA abundance, suggesting that additional regulatory mechanisms remain to be uncovered^7^. Alternative RNA splicing represents an added layer complexity not yet accounted by short read RNA sequencing.

From a single gene, through exon skipping, or intron retention, multiple transcript isoforms with distinct coding or regulatory properties are generated. More than 90 % of human genes undergo alternative splicing^8^, yielding over 100,000 transcripts from the 20,000 human genes^9^. Early studies indicate that the kidney is among the tissues with the highest splicing complexity^10^, yet splicing events and the genetic architecture of renal splicing remains largely uncharted. This knowledge gap is clinically important, as splice-altering variants can have profound biological consequences and, crucially, are amenable to therapeutic modulation, as demonstrated by the success of splice-switching oligonucleotides in neuromuscular and metabolic diseases^11,12^.

Progress in the splicing field has been constrained by technological limitations. Short-read RNA sequencing offers high depth and robust quantification of splice junction usage but struggles to reconstruct full-length transcripts when isoforms share exons. In contrast, long-read sequencing technologies such as Pacific Biosciences (PacBio) Iso-Seq can capture complete isoform structures with single-molecule resolution and high sequence accuracy^13^, although typically at lower read depth^14^. Thus, a hybrid approach combining the strengths of short- and long-read sequencing is ideally suited for building an isoform-resolved genetic map of the human kidney.

To address these gaps, we generated short-read RNA-sequencing data from 404 human kidney cortex samples, mapped cis-acting splice-QTLs (sQTLs) and eQTLs, and integrated these regulatory maps with GWAS data for CKM traits. We further applied PacBio long-read sequencing to a representative subset of samples to catalogue novel transcripts and to assign junction-level sQTL signals to full-length isoforms. This integrated framework provides the first high-resolution, isoform-resolved atlas of genetic regulation in the human kidney and establishes a foundation for mechanistic studies that connect non-coding genetic variation to CKM disease risk.

## Results

### Characterization of splicing events in the human kidney

Human kidney cortex samples from 404 nephrectomy specimens were analyzed to characterize splicing patterns and their potential relevance to kidney function. Samples were collected from four clinical sites across the United States, and the baseline clinical characteristics of the cohort are shown in Supplementary Table 1. The mean age of participants was 62.5 years, with 66 % self-identifying as male and 88 % as White; 56 % of individuals had diabetes mellitus and 73 % had essential hypertension. The mean estimated glomerular filtration rate (eGFR) was 61 mL min□¹ 1.73 m□², and its distribution was approximately normal, providing a balanced representation of both individuals with preserved kidney function and those with varying degrees of chronic kidney disease.

Alternative splicing is a post-transcriptional mechanism that generates multiple transcript isoforms from a single gene by varying the inclusion or exclusion of exonic regions at splice sites. Splicing events from short-read bulk RNA sequencing was quantified using LeafCutter^15^. Traditional RNA quantification aligns the raw reads to annotated exons and generates gene counts by normalizing to gene length and library size. Here we realigned the RNAseq reads and extracted reads that span two adjacent exons (exon-exon junctions). LeafCutter identifies splicing events based on these junction-spanning reads and organizes them into intron excision clusters (IECs). For each IEC, we calculated an intron excision ratio (IER), representing the proportion of reads supporting a specific excision event, and derived a corresponding numeric percentage reflecting the expression of individual intron excision isoforms (IEIs) (Fig. 1A). In total, 291,790 IEIs across 69,737 IECs were detected. After applying quality control filters to exclude low-coverage events, defined as absent in more than 50 % of samples, 69,403 high-confidence IEIs across 37,837 IECs were retained for downstream analysis.

**Figure 1.**
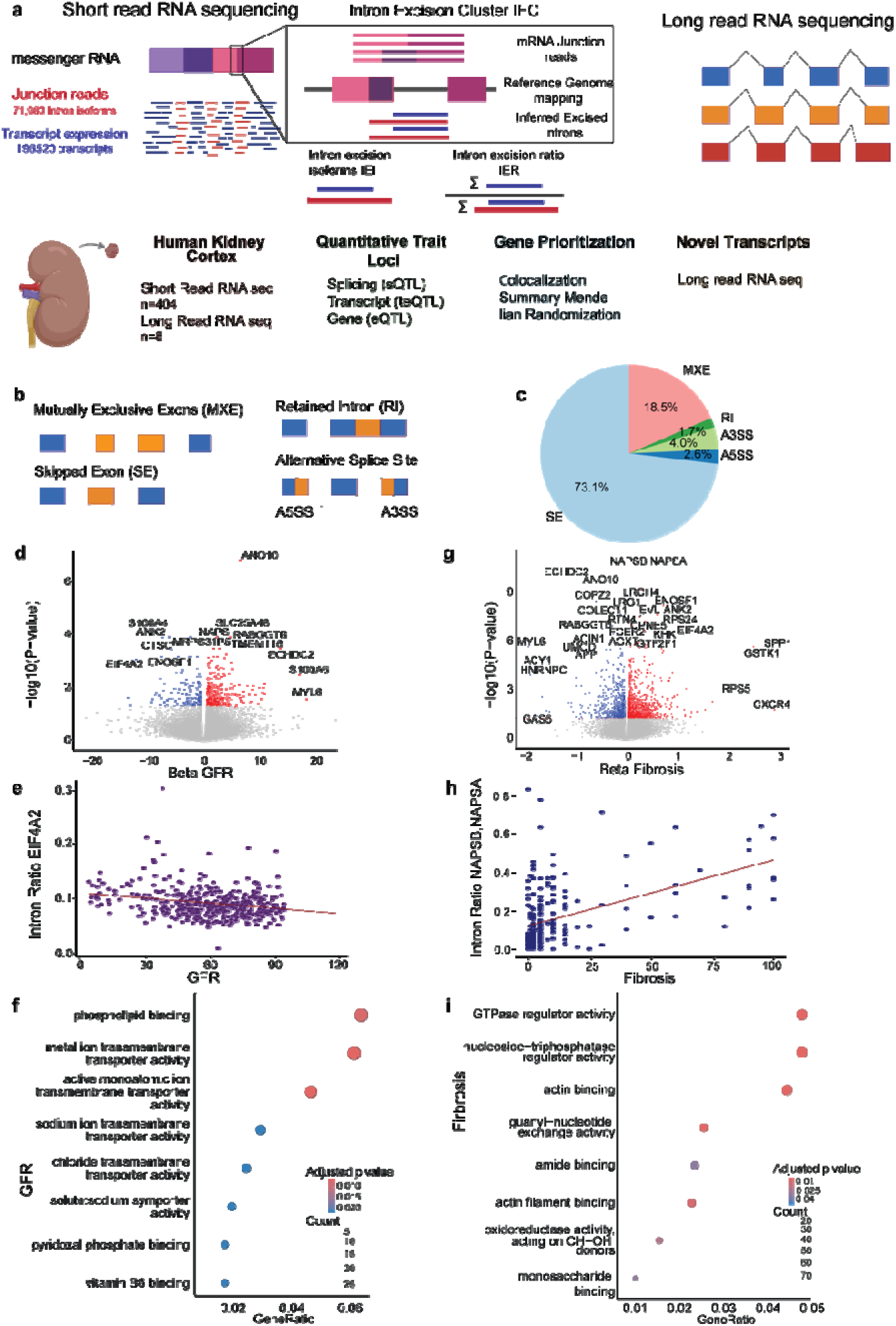
Characterization of splicing events in the human kidney. A. Junction reads from short-read RNA sequencing were used to quantify splicing events as the intron excision ratio (IER) of each intron excision isoform (IEI) within an intron excision cluster (IEC). Long-read sequencing was used to identify full-length isoforms. A schematic of the workflow is shown. Short-read sequencing was performed on 404 human kidney cortex samples, and quantitative trait locus (QTL) mapping and gene prioritization were performed using genome-wide association study (GWAS) datasets. Long-read sequencing from 8 samples was used to identify novel isoforms. B. The types of splicing events obs rved. C. The relative splicing events in the kidney dataset, with skipped exons representing the most common event. D. Volcano plot summarizing the results of linear regression between splicing events and estimated glomerular filtration rate (GFR). X-axis, the GFR and y axis is the negative logp value. E. Change in intron excision ratio of an IEI of EIF4A2, a gene with a significant association with GFR. Each dot is one human kidney sample, x-axis GFR and y-axis the intron ratio of EIF4A2. F. Gene ontology, functional pathway enrichment of gene intron excision ratio significantly associated with and GFR. G. Volcano plot showing the results of linear regression between splicing events and interstitial fibrosis. X-axis, the relative fibrosis score and y axis is the negative logp value H. Change in intron excision ratio of an IEI of NAPSA/NAPSB, a gene significantly associated with interstitial fibrosis. Each dot is one human kidney sample, x-axis GFR and y-axis the intron ratio of NAPSA. I. Gene ontology, functional pathway enrichment of genes significantly associated with intron excision ratio and interstitial fibrosis.

To quantify splicing complexity, we measured intron excision events using LeafCutter-defined intron excision clusters (IECs). Across the cohort, the median number of IECs per gene was 2 (interquartile range [IQR] = 3), and each IEC contained a median of 3 intron excision isoforms (IEIs) (IQR = 3) (Supplementary Fig. 1A–B). Exon skipping occurs when a cassette exon is spliced out of the mature transcript, whereas in intron retention, the intron is retained in the mature transcript (Fig. 1B). In the human kidney cortex, exon skipping accounted for 73.1% of all identified splicing events (Fig. 1C), consistent with patterns observed across other human tissues^16^. The next common event was a mutually exclusive exon representing 18% of all identified events. The number of isoforms per gene increased with the number of IECs, indicating that genes with more alternative splicing events exhibited greater transcript diversity. These patterns were consistent with observations in other tissues (Supplementary Fig. 1B–D).

To examine the potential clinical relevance of these splicing events, we performed linear regression analyses assessing the association of isoform usage to estimated glomerular filtration rate (eGFR), adjusting for age and sex as covariates. Intron excision ratios (IER) were arcsine and square-root transformed to stabilize variance. These analyses identified 496 genes with isoforms associated with eGFR (at FDR< 0.05) (Fig. 1D). Functional pathway enrichment analyses showed that genes associated with eGFR were enriched for membrane transport and ion handling pathways, which is the main function of the kidney tubule cells (Fig. 1F). As an example, we observed a strong correlation between the eukaryotic elongation factor family gene (EIF4A2) intron excision ratio with GFR (Fig. 1E) EIF4A2^17^. We next looked for splicing changes associated with structural damage in the kidney defined by interstitial fibrosis (log-transformed interstitial fibrosis scores was available for 225 samples). Similarly, 1875 genes have isoforms associated with interstitial fibrosis (Fig. 1G). As an example, an aspartic protease, *NAPSA/NAPSB*^18^ correlated with interstitial fibrosis (Fig. 1H). Functional pathway enrichment analysis of these genes include actin-cytoskeleton and metabolic pathways (Fig. 1I). These findings suggest a widespread shifts in alternative splicing during disease.

### Long Read Sequencing identifies novel isoforms

Typical short-read sequencing is limited to span of around 150 nucleotides; therefore, it can resolve the inclusion or exclusion of single exons but is unable to reconstruct full-length transcript isoforms. To overcome this limitation, we performed long-read RNA sequencing on eight human kidney cortex samples using the PacBio platform (Methods). Full-length transcripts were identified using the IsoSeq pipeline and filtered to remove artifacts, low-coverage isoforms, and transcripts with non-canonical splice junctions. Most transcripts detected were novel: 61.4% of isoforms identified had not been previously annotated. In total, 73,866 reference-annotated transcripts were recovered, while novel transcripts were further classified into 34.7% novel not-in-catalog (NNC) isoforms and 26.7% novel in-catalog (NIC) isoforms (Fig. 2A). Similar to other organs, the median transcript length across all isoforms was 2,030 nucleotides^19^ (Supplementary Fig. 2A), and there was no significant difference in length distribution between annotated and novel isoforms (Supplementary Fig. 2B), indicating that the newly detected isoforms are comparable in size to previously known transcripts.

**Figure 2.**
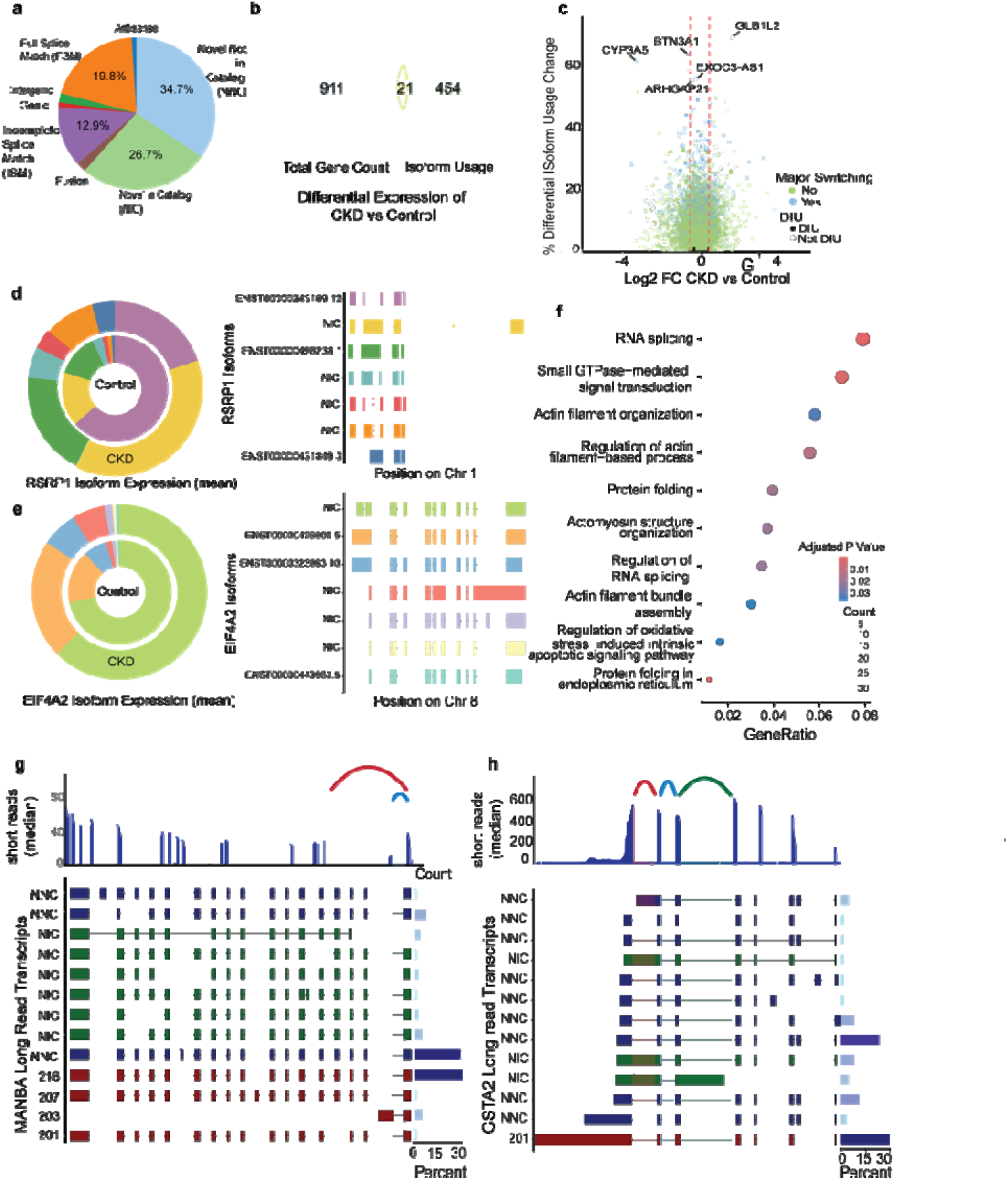
Long Read Sequencing identifies novel isoforms. A. Distribution of isoforms identified by long read RNA sequencing. B. Differential gene and isoform usage between CKD and control samples. C. Plot showing genes with significant differential isoform usage. X-axis fold change difference between controls and disease, y-axis differential isoform usage. Genes with a switch in the major isoform are colored in blue. D. Difference in mean expression between control and CKD of RSRP1 gene showing class switching. The left panel shows the long read sequencing resolved isoforms. The right panel shows distribution of isoforms in healthy and diseased kidneys. E. Difference in mean expression between control and CKD of EIF4A2 that was also demonstrated by short read sequencing. The left panel shows the long read sequencing resolved isoforms. The right panel shows distribution of isoforms in healthy and diseased kidneys. F. Gene ontology, pathway enrichment of genes with significant DIU. G. MANBA gene. Sashimi plot shows the median short reads that were sequenced. Chromosomal location and each exonic reads. Under all MANBA isoforms (known and novel) and the right panel indicates the detected relative isoform amount. 9 novel transcripts were identified and 4 annotated transcripts. An NNC transcript has the second highest abundance. H. GSTA2 gene. Chromosomal location and each exonic reads. Under all GSTA2 isoforms (known and novel) and the right panel indicates the detected relative isoform amount. There is only one known transcript of this gene. 12 novel transcripts were identified and with a relatively high abundance.

To investigate how full-length transcript isoforms differ between CKD and control kidneys, we performed a differential isoform usage (DIU) analysis using tappAS. Unlike traditional differential gene expression, which measures changes in total expression, DIU assesses whether the relative abundance of transcript isoforms from the same gene changes between conditions. We identified 475 genes with significant (FDR<0.05) changes in isoform usage in CKD (Figure 2B, C). Among these, 57 genes exhibited a major isoform switch, where the most abundantly expressed isoform in control samples was replaced by a different dominant isoform in CKD. In parallel, standard differential gene expression analysis identified 932 genes associated with CKD (FDR<0.05); however, only 21 genes overlapped with those identified by DIU (Fig. 2B), indicating that many transcript-level changes occur independently of total gene expression differences. Of the DIU genes, 40 had been linked to estimated glomerular filtration rate (eGFR) in short-read RNA sequencing. (Supplementary Fig. 2C). Notably, the splicing regulator RSRP1 showed a shift in predominant isoform expression from the annotated transcript ENST00000243189.12 to a novel in-catalog (NIC) transcript in CKD (Fig. 2D), and EIF4A2 also demonstrated significant changes in isoform usage (Fig. 2E). Gene Ontology enrichment analysis of DIU genes revealed significant overrepresentation of pathways related to splicing mechanism, cellular regulation, subcellular organization, and protein localization (Fig. 2F). These results highlight the value of isoform-level analysis in uncovering transcriptional alterations that are not readily detectable in gene-level approaches.

To further evaluate the relationship between long-read and short-read data, we compared isoforms identified from long-read sequencing to splice junctions detected in short-read RNA sequencing. Sashimi plots were used on short read data to plot the median read count for each exon. The long-read data provided full-length transcript context, clarifying how exon– exon junctions observed in short-read data map to specific transcript structures. For example, in the beta mannosidase gene (MANBA), the junction spanning chr4:102753995–102760718 (hg19 coordinates chr4:103675153–103681875) corresponded uniquely to the full-length transcript MANBA-203 (Fig. 2G). This enables isoform-specific quantification from short-read data, as this junction distinguishes MANBA-203 from all other isoforms and can be visualized through sashimi plots, where read density over the unique exon provides transcript-level resolution. In contrast, the GSTA2 gene highlighted the limitations of short-read approaches: multiple GSTA2 isoforms share common exon–exon junctions, making it difficult to resolve which full-length transcripts were expressed. Furthermore, GSTA2 has only one annotated reference transcript, suggesting that conventional short-read analysis may substantially underestimate its isoform diversity (Fig. 2H).

The long-read sequencing data reveals a remarkable abundance of previously unannotated isoforms. Together, these findings underscore the complexity of kidney transcriptomes and highlight the critical value of long-read sequencing for achieving isoform-level resolution in human tissues.

### Splicing events associated with genetic variations

To begin differentiating between splicing events causing kidney disease vs those that are the consequence of kidney disease, we performed splicing quantitative trait locus (sQTL) mapping by defining the association between genotype variants and intron excision measurements from short-read RNA-seq (Fig 3A). In parallel, we generated matched cis-QTL maps for total gene expression (eQTL) and full-length transcript expression using annotated reference transcripts from Gencode v19^20^ (teQTL). A gene with any significant intron exclusion or transcript QTL at a qvalue of 0.05 was considered a significant sQTL or teQTL gene respectively. Although both sQTL and teQTL analyses capture information on spliced isoforms, they did so through different strategies: sQTLs quantify splicing by modeling intron excision, whereas teQTLs infer transcript usage by mapping the reads to the known full-length transcripts. All QTL analyses were conducted within a ±1 Mb window around each gene and adjusted for confounders, including age, sex, batch, site, genotype-derived principal components, and PEER factors. Statistical significance was determined using permutation testing and Benjamini–Hochberg correction to control the gene-level false discovery rate at 5% (Methods). The whole genome sequencing genetic data underwent alignment and QC.

**Figure 3.**
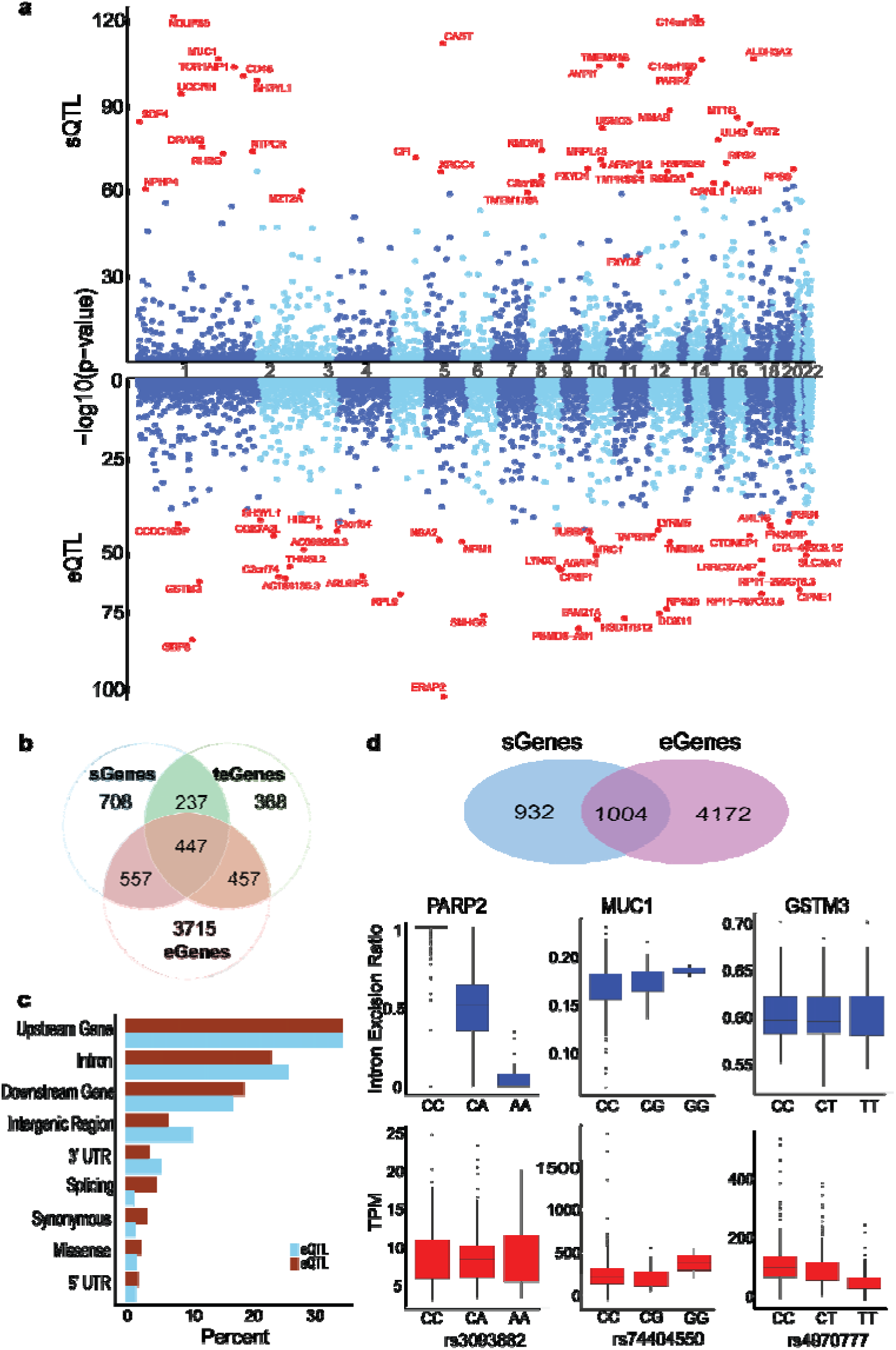
Splicing events associated with genetic variations. A. Manhattan plot of sQTL and eQTL. The top significant genes in each dataset is highlighted in red. X-axis chromosomal location, y-axis log(p) value. B. Overlap of sQTL genes, teQTL genes and eQTL genes. A total of 1948 sQTL genes, 1509 teQTL genes and 5314 eQTL genes were identified, with 452 sQTL genes are also eQTL genes and have a significant transcript that is a teQTL gene. C. The functional annotation of sQTL (brown) and eQTL (blue) variants. D. Difference in intron excision ratio and RNA expression (TPM) by allele dose of the top SNP in PARP2 (sQTL gene only), MUC1 (sQTL gene and eQTL gene) and GSTM3 (eQTL gene only). The x-axis is the reference and alternate allele of the SNP and y-axis is the relative expression (TPM or IER) of the gene.

Our sQTL analysis identified 1,948 genes with significant (FDR<0.05) sQTLs (sGenes), 5,176 genes with significant (FDR<0.05) eQTLs (eGenes), and 1,502 genes with significant (FDR<0.05) teQTLs (teGenes) (Fig. 3B). Despite their shared focus on isoforms, only 35% of sQTL genes overlapped with teQTL genes, suggesting that transcript-level approaches miss a substantial proportion of splicing variation. In contrast, 60% of teQTL genes overlapped with eQTL genes, whereas only 52% of sQTL genes did, highlighting the partial independence of splicing and expression regulation (Fig 3B). Furthermore, only 25% of sQTL genes overlapped with eQTL genes, underscoring the value of isoform-level analyses in identifying regulatory events that would be missed by conventional gene-level approaches.

We next compared our sQTL dataset to external kidney sQTL sources. Of the 1,948 sQTL genes, 66.7% overlapped with those reported in GTEx v8, which includes sQTLs from 72 kidney cortex samples (Supplementary Fig. 3A). The effect sizes (β coefficients) for overlapping sQTLs were highly concordant, with a Pearson correlation of 0.937 (Supplementary Fig. 3B). Similarly, 69.9% of sQTL genes identified in our analysis were also found in another human kidney study^21^, confirming the robustness and reproducibility of our results.

To further characterize sQTL and eQTL regulatory architecture, we examined the genomic locations of lead variants. Compared to lead eQTL variants (eSNPs), which had a median distance of 120 base pairs from the transcription start site (TSS), sQTL variants (sSNPs) localized farther away, with a median distance of 722 base pairs. We annotated these variants using ChromHMM chromatin states and kidney-specific epigenomic datasets including ATAC-seq and Histone modifications (Methods). sSNPs in open chromatin regions were more frequently enriched in transcription state marking the bodies of actively transcribed genes, whereas eSNPs tended to localize to the transcription start site and repressors (Supplementary Table19). Functional annotation using SNPeff revealed that a higher proportion of sSNPs were predicted to directly impact splicing—most commonly through intronic, splice donor/acceptor, or splice region variants—though the overall distributions of predicted consequences for sSNPs and eSNPs were not significantly different (chisquare pvalue 0.2) (Fig. 3C).

To evaluate the functional consequences of sQTLs versus eQTLs, we performed gene ontology enrichment analyses. sQTL genes were significantly enriched for pathways related to RNA processing, ion transport, and epithelial differentiation, whereas eQTL genes were more strongly associated with transcriptional regulation and metabolic processes. Shared enrichment was observed in oxidoreductase activity pathways (Supplementary Fig. 2D). Our analysis indicated that sQTL, eQTL and teQTL data sometimes can be concordant for example at Mucin 1 (*MUC1*) gene, where the changes in splicing event can also be identified at eQTL level. At many cases the eQTL aggregation missed sQTL information such as the case of poly(ADP-ribosyl)transferase-like 2 protein (*PARP2*). While Glutathione S-Transferase Mu 3 protein (*GSTM3)*, there is a true eQTL effect without detectable changes in splice variants (Fig. 3D). These examples highlight how genetic variants can exert regulatory effects through distinct mechanisms—modulating splicing, expression, or both—and illustrate the value of examining individual loci to disentangle the specific contributions of each regulatory pathway.

### Colocalization to nominate Genes with significant associated with eGFR

To understand whether splicing changes might mediate the effect of genetic variants on kidney function, we integrated our splicing QTL (sQTL) summary statistics with a genome-wide association study (GWAS) of estimated glomerular filtration rate (eGFR) conducted in 1.77 million individuals of European ancestry^22^ (Methods). We applied Bayesian colocalization to identify loci where the same genetic variant likely drives both splicing variation and eGFR association. Genes with a posterior probability of colocalization (PP4) ≥ 0.80 were prioritized to harbor kidney-relevant sQTLs. This analysis identified 124 sQTL genes with at least one significant intron excision event that colocalized with an eGFR GWAS locus (Fig. 4A, Supplementary Table 5). These include CHMP1A, MANBA, ACTN4, ACSM3, and MGMT, where the associated splice events may provide a mechanistic explanation for how noncoding genetic variants influence kidney disease development through altered isoform composition.

**Figure 4.**
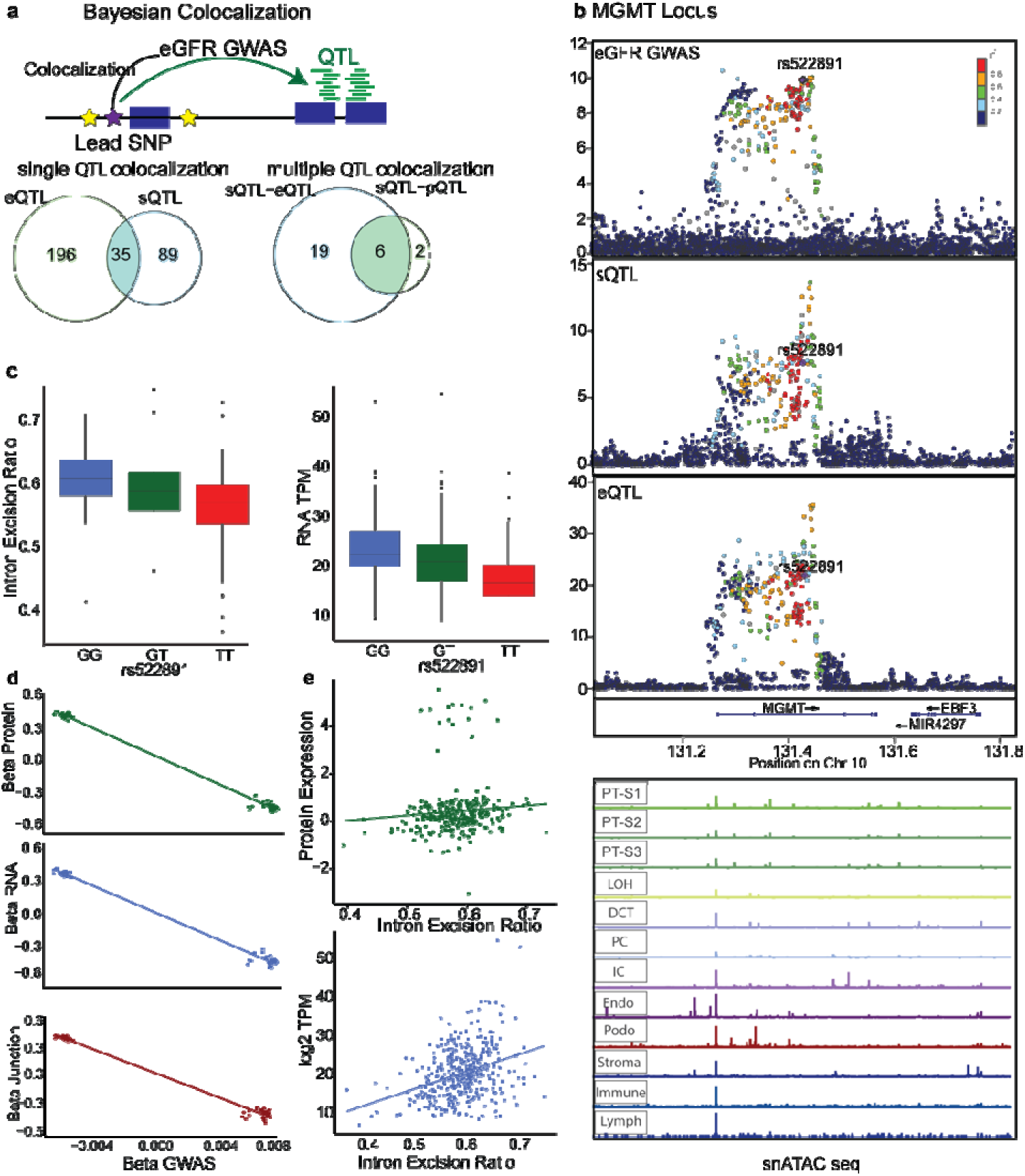
Colocalization to nominate Genes with significant associated with eGFR. A. Colocalization of human kidney sQTL with eGFR GWAS dataset using 1.77 million subjects of European ancestry. COLOC was used for colocalization with one QTL trait and MOLOC for multitrait colocalization with eGFR GWAS. 124 sQTL genes and 231 eQTL genes had SNPs that colocalized with index SNPs in the eGFR GWAS. 35 of these genes overlapped. Using MOLOC, eQTL and pQTL was analyzed in turn with sQTL for multitrait colocalization with eGFR GWAS. 25 sQTL genes a d eQTL genes and 8 sQTL genes and pGenes colocalized with eGFR GWAS. 6 genes overlapped between both analyses that includes MGMT gene. B. Locus Zoom plot of MGMT locus in the GWAS, sQTL and eQTL dataset. snATAC seq of this locus is also shown. X-axis chromosomal location, y-axis is the strength of the association -log(p) value. C. intron excision ratio of the 131334641:131506159 junction and TPM of MGMT by allele dose of rs522891. X-axis is the genotype, y-xis is the relative expression (TPM) or excision ration (IER). D. Beta values of the MGMT locus comparing the Beta of GWAS SNPs on Splicing, RNA and Protein. E. Change in RNA expression (TPM) and protein expression (log2 protein expression) by IER of the 131334641:131506159 junction. The x-axis shows the GWAS effect sizes while the y-axis relative gene or protein expression or splicing.

For comparison, colocalization of expression QTLs (eQTLs) with eGFR GWAS loci identified 231 eQTL genes (Fig. 4A, Supplementary Table 6), including established kidney-relevant genes such as UMOD, DPEP1, SH3Y1, CISD2, and TRIM6. Notably, 5.2% of eGFR-associated loci colocalized with sQTLs but not eQTLs, suggesting that alternative splicing may represent the primary regulatory mechanism for these genetic signatures. Thirty-five genes colocalized in both the sQTL and eQTL datasets, including RNF212, CHMP1A, MANBA, DEPDC5, and ACSM3. At these loci, a single genetic variant appears to influence both splicing and overall gene expression, suggesting the underlying splicing change is responsible for the change in gene expression. This convergence of molecular effects strengthens the case for causality, as variants that affect both transcript abundance and isoform structure may have a greater functional impact— such as promoting expression of a disease-associated isoform or altering the balance between functional and non-functional transcripts.

To determine whether splicing and expression QTLs at the same gene are driven by a shared causal variant, we applied multi-trait colocalization using MOLOC—a Bayesian framework that tests whether associations across multiple molecular phenotypes arise from a single underlying variant rather than from distinct, nearby signals¹□. This analysis revealed 25 genes in which sQTL and eQTL signals colocalized, indicating that the same genetic variant likely regulates both splicing and overall transcript abundance (Supplementary Table 7).

We next compared genetic effects on splicing and total RNA with proteoform-level quantitative trait loci (pQTLs). Proteoform profiles from 325 human kidney cortices, generated by SomaScan, yielded 873 significant pQTLs^23^. Colocalization analysis showed that eight splicing QTL (sQTL) genes and eleven expression QTL (eQTL) genes share the same causal variants with pQTLs. Twenty-one genes appeared in all three datasets. When we applied the MOLOC framework to integrate sQTL–eQTL–pQTL signals, six genes (MGMT, CHMP1A, NT5DC2, MANBA, GSTA1, and the antisense transcript GSTA2-AS1) displayed a single variant that simultaneously influences splice choice, steady-state RNA abundance, and protein concentration. This multi-layer convergence, together with genome-wide significant eGFR associations at the same loci, provides strong causal evidence for these genes in kidney function. The causal role of CHMP1A, MANBA and GSTA in kidney function regulation has already been supported by cell and animal model experiments^24–26^.

MGMT (O6-methylguanine-DNA methyltransferase) emerged as a compelling new candidate gene. MGMT encodes a DNA repair enzyme known for its role in protecting against genotoxic damage and modulating chemotherapy response^27^. In our study, a common variant at the MGMT locus colocalized with sQTL, eQTL, and pQTL signals, as well as with a significant GWAS locus for eGFR (Fig. 4B). This suggests that a single genetic variant may coordinately regulate MGMT splicing, transcript abundance, and protein levels, thereby influencing kidney function.

To assess the reproducibility of this association, we examined multiple external datasets. The same MGMT intron excision event was identified as an sQTL in GTEx kidney cortex samples^28^ and in an independent biopsy-based kidney QTL study^21^. The MGMT eQTL was also confirmed by prior kidney eQTL studies^29,28,30^. Among multiple MGMT intron excision ratios (IERs), one splice junction—chr10:129536377–129707895 (hg19 coordinates: chr10:131334642– 131506159)—was markedly associated with the eGFR GWAS lead SNP rs522891. The risk allele at this SNP was associated with reduced intron excision (lower IER) and lower MGMT expression (Fig. 4C). These regulatory effects were directionally consistent with the GWAS signal: greater MGMT expression and higher IER were associated with lower eGFR, implicating MGMT as a risk gene for reduced kidney function (Fig. 4D). Furthermore, higher IER at this junction correlated with increased MGMT RNA and protein levels (Fig. 4E), supporting a functional link between splicing and protein abundance. snATAC-seq revealed open chromatin around the TSS in all cell types; however, the strongest GWAS and QTL signals overlapped accessible regions within the gene body, predominantly in endothelial and podocyte cells.

To place the splice event in the context of MGMT isoform structure, we analyzed annotated and novel isoforms. Among the three annotated transcripts, only one (MGMT-203) lacks the disease-associated exon-exon junction and accounts for a minor fraction of total expression (Extended Data 2). Long-read RNA sequencing identified seven additional MGMT isoforms, two of which contain the relevant intron, while five do not. Although the splicing event cannot be uniquely assigned to a single full-length isoform, its presence in most major transcripts, its reproducibility across datasets, and its association with expression, protein level, and disease risk collectively support its functional relevance.

Together, these findings establish MGMT as a genetically regulated, multi-layered effector of kidney function. The convergence of splicing, expression, and protein regulation at a single locus that also harbors a GWAS signal provides compelling evidence that MGMT contributes to kidney disease risk through coordinated regulatory mechanisms.

### Mendelian Randomization to identify causal genes

While colocalization analysis provides strong evidence that the same variant underlies both a molecular QTL (e.g., sQTL or eQTL) and a GWAS signal for eGFR, it does not establish a directional causal relationship—namely, whether the molecular trait (e.g., altered splicing or gene expression) mediates the genetic effect on disease, or is simply correlated due to linkage disequilibrium. To address this, we applied Summary Mendelian Randomization (SMR), a statistical approach that tests whether genetically predicted variation in a molecular trait causally influences the complex trait. SMR integrates QTL summary statistics and GWAS results within an instrumental variable framework, using the top QTL-associated SNP as a proxy for the molecular trait to estimate its effect on eGFR. To reduce the risk of horizontal pleiotropy—where the same variant influences multiple traits independently rather than through a shared mechanism—we additionally applied the HEIDI (Heterogeneity in Dependent Instruments) test. The HEIDI test helps distinguish true causal mediation from confounding due to linkage disequilibrium LD.

Using this two-step framework, we identified 64 sQTL genes and 87 eQTL genes with evidence of causally influencing eGFR (Fig. 5A, Supplementary Tables 9 & 10). Significance was defined using a Bonferroni-adjusted SMR p-value < 0.05 and a HEIDI p-value < 0.01. Among these, 34 sQTL genes and 40 eQTL genes were also supported by colocalization analysis, increasing confidence in their role as kidney disease susceptibility genes. Notably, 21 sQTL genes were uniquely identified through splicing data, underscoring the added value of isoform-level analyses in uncovering regulatory mechanisms not captured by gene-level expression. The top 10 prioritized genes are shown in Table 1.

**Figure 5.**
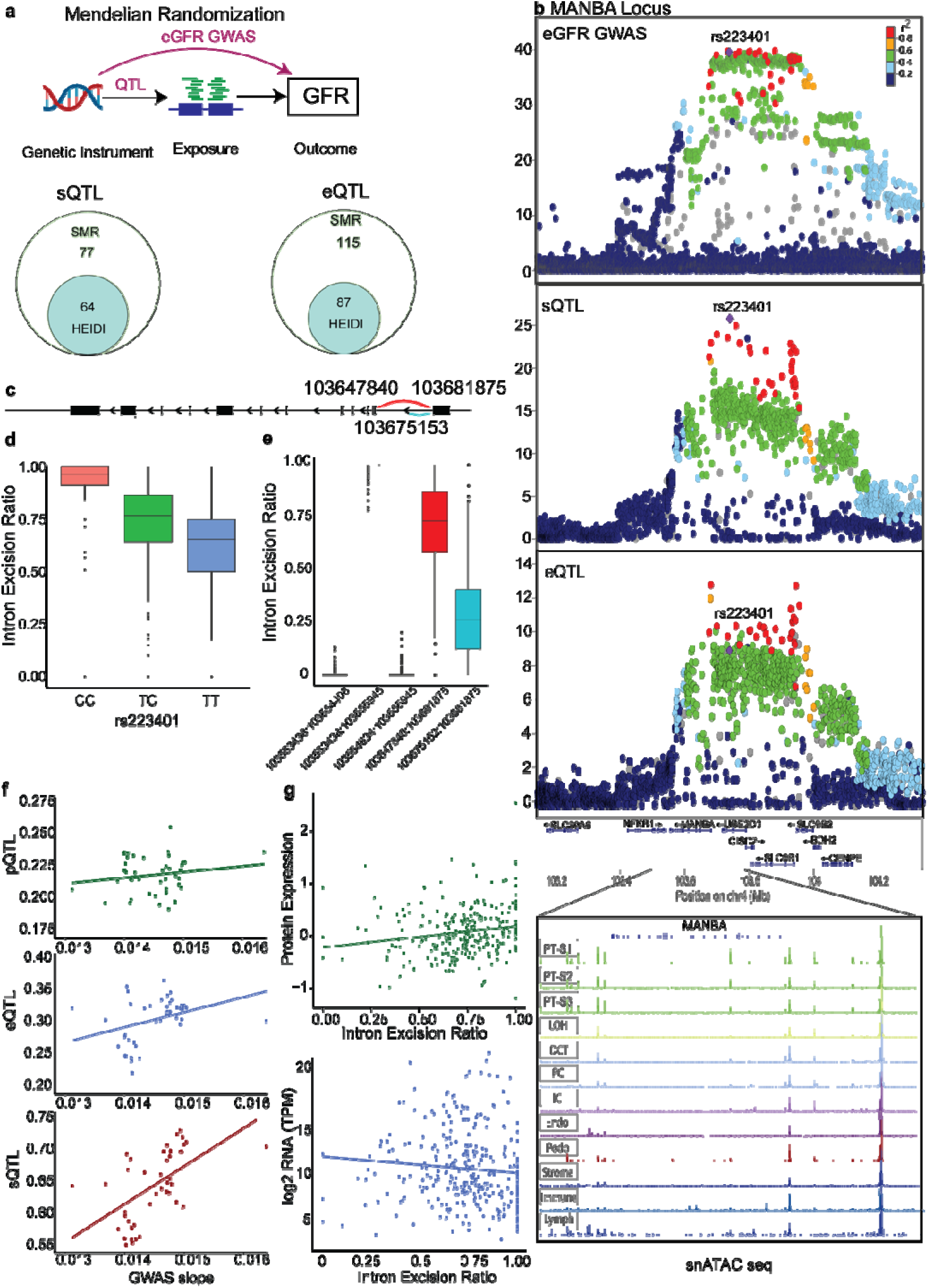
Mendelian Randomization to identify causal genes. A. Mendelian Randomization of eGFR GWAS with sQTL and eQTL data. eGFR was used as the outcome using the results of the eGFR GWAS. sQTL and eQTL was used as the exposure to prioritize causal genes. The results of each outcome are displayed, and the overlap of the prioritized genes were compared with the COLOC results. 34 sQTL genes and 40 eQTL genes were considered as causally associated with eGFR. B. Locus Zoom plot of the MANBA locus in GWAS, sQTL and eQTL. snATAC seq of this locus is also shown. X-axis is chromosomal location y-axis is strength of association -log(p) value. Followed by human kidney single nucleus ATAC-seq tracks. C. Intron excision ratio of each intron isoform in intron clusters mapped to the MANBA gene. Only events in one cluster showed significant variation among the cohort. D. IER by allele dose of rs223401 SN 103647840:103681875 junction showing a decrease in IER (y-axis) with the alternative allele. E. Plot of for the the beta coefficient of overlapping SNPs in the MANBA Locus comparing GWAS beta value with sQTL, eQTL and pQTL. SNPs with a positive GFR effect are associated with a higher IER at the 103647840:103681875 junction, RNA and protein expression F. Change in RNA expression (TPM) and protein expression (log2 protein expression) by IER of the 103647840:103681875 junction. This shows a weak correlation of the IER and RNA or protein expression.

**Table 1.** Top 10 significant genes nominated in both the sQTL and eQTL dataset using COLOC/SMR with eGFR GWAS. PP H4 (posterior probability of H4 by COLOC), b_SMR (beta coefficient of SMR), p_SMR (p-value of SMR), p_HEIDI (p value of HEIDI test)

| Gene | Junction | sQTL |  |  |  | eQTL |  |  |  |
| --- | --- | --- | --- | --- | --- | --- | --- | --- | --- |
|  |  | PP H4 | b_SM R | p_SMR | p_HE IDI | PP H4 | b_SM R | p_SMR | p_HEIDI |
| TCEA3 | chr1:23707967:23710839 | 0.964 | -0.021 | 3.15E-08 | 0.097 | 0.969 | -0.074 | 7.15E-08 | 0.145 |
| METTL10 | chr10:126454177:126463244 | 0.969 | 0.021 | 6.38E-12 | 0.101 | 0.998 | 0.032 | 2.27E-10 | 0.249 |
| MGMT | chr10:131334641:131506159 | 0.982 | -0.014 | 9.96E-07 | 0.191 | 0.995 | -0.015 | 8.60E-09 | 0.491 |
| UMOD | chr16:20360534:20361972 | 0.812 | -0.114 | 2.45E-10 | 0.35 | 1 | -0.291 | 1.11E-14 | 0.626 |
| ACSM3 | chr16:20791229:20792036 | 0.988 | -0.016 | 9.47E-06 | 0.053 | 0.98 | 0.031 | 1.83E-05 | 0.38 |
| CHMP1A | chr16:89713122:89713611 | 0.993 | 0.037 | 3.65E-08 | 0.034 | 1 | 0.054 | 4.92E-12 | 0.028 |
| SH3YL1 | chr2:219001:229966 | 0.978 | -0.007 | 2.90E-13 | 0.593 | 1 | 0.019 | 1.48E-12 | 0.115 |
| CPNE1 | chr20:34214303:34214657 | 0.814 | -0.01 | 1.93E-06 | 0.057 | 0.986 | 0.008 | 4.18E-08 | 0.3 |
| MANBA | chr4:103647840:103681875 | 0.991 | 0.02 | 9.86E-17 | 0.357 | 1 | 0.039 | 3.06E-10 | 0.136 |
| GSTA1 | chr6:52615497:52616375 | 0.973 | 0.023 | 1.97E-07 | 0.294 | 0.987 | -0.067 | 1.53E-09 | 0.061 |

One particularly compelling example is MANBA (Beta Mannosidase), a lysosomal enzyme. MANBA was identified as both an sQTL and eQTL gene and passed colocalization, SMR, and HEIDI filtering with the eGFR GWAS (Fig. 5B). snATACseq showed the lead variants overlay open chromatin in all cell types however, the strongest GWAS and QTL signals overlapped accessible regions within the gene body in predominantly proximal tubule cells (Fig. 5B). While previous studies have implicated MANBA in kidney disease through expression and protein associations^24,31^, the role of alternative splicing had not been explored. Our findings suggest that specific MANBA splice isoforms, rather than total gene expression alone, mediate its genetic effect on kidney function.

Both intron isoforms within a single MANBA cluster were supported by SMR and COLOC, and MOLOC analysis indicated that splicing, RNA expression, and protein abundance at this locus are driven by the same causal variant. These intron isoforms showed complementary expression patterns (Fig. 5E), and the associated SNPs had opposing effect directions on eGFR, suggesting that the relative abundance of each isoform may determine the gene’s overall impact on kidney function. Specifically, similar to the role of the total gene, a higher expression of the isoform spanning chr4:102726683–102760718 (hg19: chr4:103647841–103681875) was associated with higher eGFR (Fig. 5F), consistent with a protective role. However, no clear correlation was observed between isoform-level intron excision ratios (IERs) and total RNA or protein abundance (Fig. 5G), suggesting that splicing may influence protein function or localization independently of overall expression level.

Long-read sequencing (Fig. 2G) revealed that only the annotated transcript MANBA-203 contains the significant intron isoform chr4:102753995–102760718 (hg19: chr4:103675153–103681875). This isoform is unusually short, comprising just two exons, and lacks a known open reading frame or proteoform^32^. These observations suggest that the disease-associated genetic variant promotes expression of a non-functional isoform, potentially leading to loss of MANBA function via isoform switching. In this model, an increase in MANBA-203 may displace functional transcripts, thus reducing effective lysosomal enzyme activity and contributing to disease risk.

Despite its genetic association, MANBA-203 accounted for only 5.1% of MANBA transcripts in our long-read dataset (Fig. 2G), and its unique exon shows low read coverage in sashimi plots, consistent with limited expression under baseline conditions. Nevertheless, genotype-specific upregulation of this isoform could exert a dominant-negative or competitive effect.

The disease-associated splicing signal at MANBA was also observed in independent datasets^21^, the same intron isoform was associated with blood pressure GWAS loci, highlighting its broader relevance to cardio-renal traits. In the GTEx kidney cortex dataset^28^, MANBA was again identified as an sQTL gene, though a different intron (chr4:102669049– 102671281; hg19: chr4:103590206–103592438) reached significance. This intron is present in all long MANBA transcripts except MANBA-203, suggesting a consistent splicing pattern in which MANBA-203 is structurally and functionally distinct from predominant protein-coding isoforms.

Together, these findings demonstrate the value of integrating colocalization, Mendelian randomization, and isoform-resolved splicing data to move beyond association and nominate genes with a likely causal role in kidney disease. The MANBA locus exemplifies how alternative splicing can fine-tune gene function and disease risk and underscores the importance of incorporating transcript-level resolution in efforts to uncover pathogenic mechanisms.

### Kidney splicing regulation links kidney function to cardiometabolic traits

To investigate whether kidney-specific regulatory variation contributes to broader cardiometabolic phenotypes, we systematically assessed the impact of kidney-derived splicing and expression QTLs (sQTLs and eQTLs) on a spectrum of cardiovascular, kidney, and metabolic (CKM) traits. These traits, now collectively recognized under the CKM syndrome umbrella, reflect the shared pathophysiologic basis of metabolic dysfunction, cardiovascular disease, and kidney injury. We hypothesized that splicing-level regulation in the kidney may underlie shared genetic architecture across CKM traits. Using colocalization analysis, we tested kidney sQTL and eQTL data against 11 CKM traits, including lipid levels (HDL, LDL, triglycerides), cardiovascular traits (systolic/diastolic blood pressure, coronary artery disease, heart failure), and metabolic traits (type 1 and type 2 diabetes, obesity). GFR, lipid traits, and blood pressure traits yielded the largest number of colocalized genes (Figure 6A), with splicing events most strongly associated with GFR, followed by HDL (48 genes) and triglycerides. Summary-based Mendelian randomization (SMR) analyses confirmed this trend, identifying 137 genes for GFR, 68 for blood pressure traits, and 122 for lipid traits (Extended Data 4).

**Figure 6.**
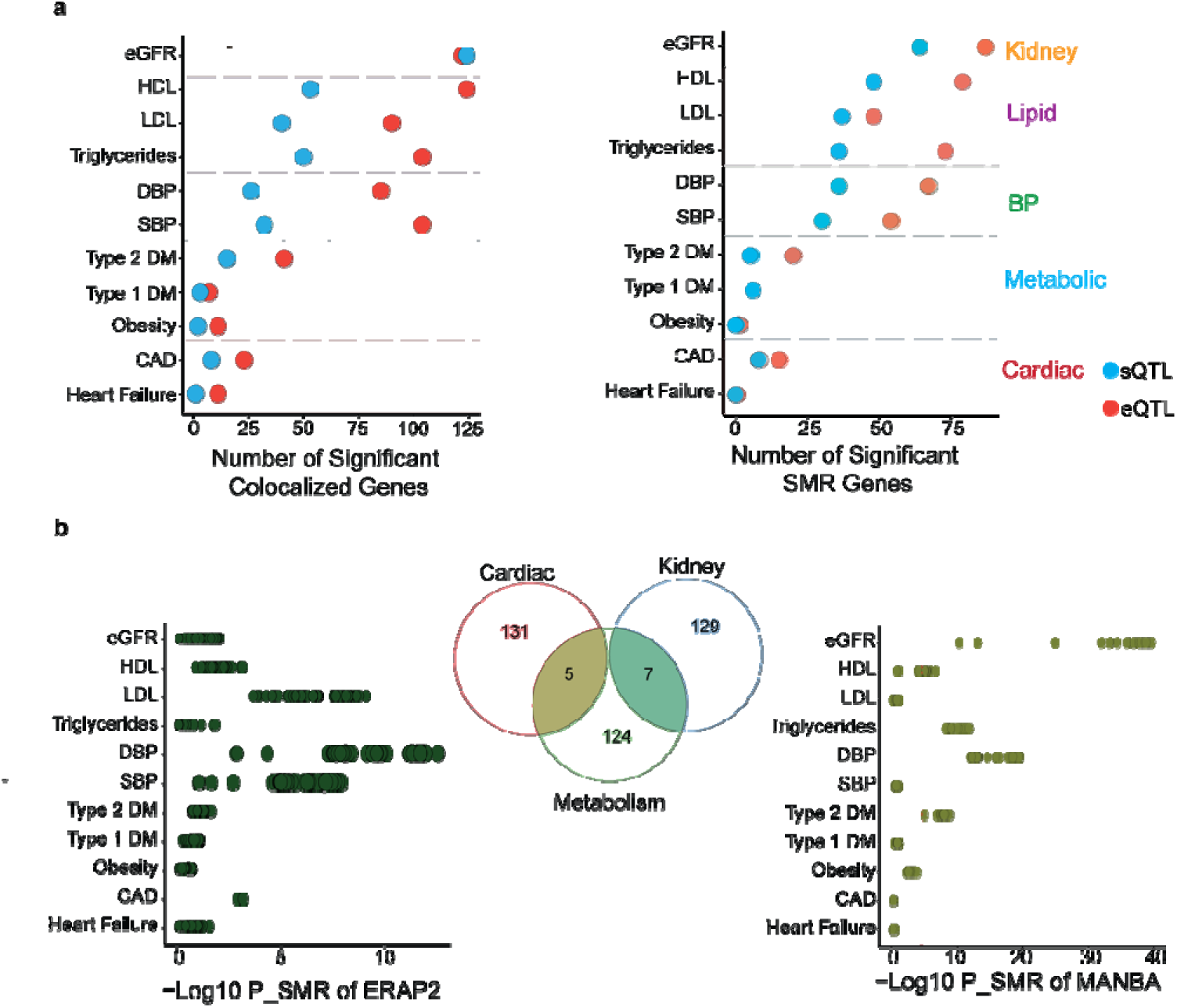
Kidney splicing regulation links kidney function to cardiometabolic traits. A. The x-axis is the number of significant colocalized genes (eQTL) in red and splicing events (blue), the y-axis is the trait group by disease categories. The x-axis is the number of significant SMR genes (eQTL) in red and splicing events (blue), the y-axis is the trait group by disease categories. B. Overlap of genes nominated by COLOC and SMR. P value (-Log10 transformed) of SNPs for each CKM trait for ERAP2 (nominated in cardiac and metabolic traits) and MANBA (nominated in GFR and metabolic traits) are shown. eGFR estimated Glomerular Filtration Rate, HDL high density lipoprotein, DBP diastolic blood pressure, DM diabetes mellitus, CAD coronary artery disease, ERAP2 Endoplasmic Reticulum Aminopeptidase 2, MANBA Beta mannosidase

We grouped traits into kidney, cardiovascular, and metabolic categories and nominated genes significant in both colocalization and SMR (Figure 6B). Seven genes—*MANBA, GSTA1, GSTA2, NT5DC2, SH3YL1, TCEA3,* and *TMEM116*—showed evidence of shared genetic regulation between kidney and metabolic traits, primarily involving lipids. Notably, *MANBA* was also significantly associated with HDL and triglycerides in SMR (Figure 6B) and with diastolic blood pressure in SMR, though not by colocalization. *GSTA1* and *GSTA2*, members of the glutathione-S-transferase A family, are involved in detoxification processes and have been previously linked to lipid levels^33^ and altered expression in acute kidney injury^34^, though not directly to chronic kidney disease.

No genes showed shared effects between kidney and cardiovascular traits. However, five genes—*ERAP2, RNF123, THAP9-AS1, SEC31A,* and *ACMSD*—overlapped between cardiovascular and lipid traits, suggesting shared regulatory mechanisms distinct from those influencing kidney function. Endoplasmic Reticulum Aminopeptidase 2 *(ERAP2)*, a gene involved in MHC class I molecules presentation^35^, showed associations with LDL, systolic, and diastolic blood pressure, but not with GFR.

Overall, most splicing QTL-associated genes were trait-specific, indicating context- or tissue-specific regulation. These findings suggest that transcript-level regulatory variation in the kidney may contribute not only to kidney disease but also to systemic CKM phenotypes, with pleiotropic effects observed in a subset of genes. Integrating splicing information reveals regulatory mechanisms not captured by expression data alone and highlights the kidney as a relevant tissue contributing to the broader genetic architecture of CKM syndrome.

### Cellular expression of genetically prioritized CKM genes in the human kidney

To determine which nephron cell types are most relevant for genes genetically linked to chronic kidney disease and cardiometabolic traits, we assessed the expression of genes prioritized through SMR and colocalization across CKM traits—including eGFR, blood pressure, lipid levels, and diabetes. We examined the expression of these genes in our adult human kidney single-nucleus RNA-seq atlas and calculated mean expression per gene across annotated cell clusters. Standardized expression values were visualized in a heatmap (Supplementary Figure 4).

This analysis revealed that CKM-prioritized genes are expressed broadly across nephron. We observed a notable expression enrichment in proximal tubule (PT) cells, particularly for genes nominated for kidney function, blood pressure and metabolic (33%, 23% and 21% respectively). TMEM116 nominated in GFR, lipid and cardiac traits showed a dominant expression in the collecting duct. Lipid-traits associated genes showed enriched expression in endothelial cells, intercalated cells and distal tubule. Whereas metabolism-associated genes showed enriched expression in podocytes and immune cells, suggesting that distinct kidney cell types may mediate genetic susceptibility to different components of the CKM spectrum.

Together, these findings highlight the proximal tubule as a potential central site of convergence for genetic risk across CKM traits, while also pointing to additional nephron and stromal cell types that may mediate trait-specific effects. These insights underscore the value of cell-type–resolved expression profiling to inform the tissue-level interpretation of genetic risk loci.

## Discussion

Our findings highlight alternative splicing as a central and underappreciated mechanism of gene regulation in the kidney, with direct relevance to both kidney-specific function and systemic cardio-renal-metabolic phenotypes. By integrating deep short-read sequencing with full-length long-read transcriptomics, we assembled the most comprehensive human kidney transcriptome to date. The majority of isoforms identified by long-read sequencing were previously unannotated, underscoring the limitations of conventional exon-centric short-read approaches and revealing widespread transcript complexity that has remained hidden^36^. Genetic analyses further demonstrated that splicing and gene expression are frequently governed by distinct regulatory architectures: fewer than half of splice QTLs overlapped with expression QTLs^28^, and approximately 5% of eGFR-associated GWAS loci colocalized exclusively with splicing events. These results highlight isoform-level regulation as a critical and independent contributor to disease risk, positioning alternative splicing—not just total transcript abundance—as a key axis of molecular pathology in kidney and systemic disease.

Integrating splicing, expression and protein QTLs through the Bayesian MOLOC framework uncovered six loci— MGMT, CHMP1A, NT5DC2, MANBA, GSTA1 and GSTA2-AS1—where non-coding genetic variants modify exon selection, shifts steady-state RNA, and alters protein concentration. Such vertical coherence across molecular layers provides strong causal evidence, elevating these genes above the long list of statistical candidates generated by large-scale association studies. At MANBA, for example, the risk haplotype favors the short MANBA-203 isoform that lacks catalytic domains, a mechanism that reconciles earlier reports of reduced MANBA activity in fibrotic tissue with the present genetic data. Similar logic applies to MGMT, where the allele that lowers DNA-repair capacity now emerges as a plausible driver of nephron vulnerability. These mechanistic insights are difficult to obtain from conventional eQTL or TWAS experiments and highlight the power of isoform-aware designs ^37–39^.

The therapeutic implications follow naturally. Splice-switching oligonucleotides and allele-directed siRNAs are already licensed for neurological and metabolic disorders^11,12^; the kidney is an attractive next organ because intravenously delivered nucleic acids accumulate efficiently in proximal□tubule cells^40^. Our dataset specifies the exact isoform ratios and risk alleles that such molecules should target and offers protein biomarkers that can confirm on-target engagement in early-phase trials. In diagnostics, isoform-specific peptides could complement serum creatinine by reporting proximal-tubule stress before global glomerular filtration rate declines, an advance that would shift intervention to a much earlier window in disease progression.

Several limitations temper our enthusiasm. Long-read sequencing still lags behind short reads in raw accuracy and depth, restricting the resolution of splice QTL mapping^41–43^. A major issue is that we do not know which transcript isoform is translated into proteoform^44,45^. Our cohort is predominantly of European ancestry, and further work is needed to test whether the same regulatory logic holds in other populations where CKD burden is high. Single-cell long-read sequencing, combined with CRISPR base editing in human kidney organoids, should refine locus-specific mechanisms and reveal whether causal splice events are confined to particular cell types^46^.

Even with these caveats, the present study establishes isoform-level regulation as a key axis of genetic influence on kidney function and related systemic traits. Future association studies that ignore splicing risk leaving causal variants undiscovered and therapeutic avenues unexplored. By delivering a high-confidence short list of genes in which one variant orchestrates transcript structure, RNA abundance and protein output, we provide a realistic foundation for isoform-guided diagnostics and interventions in chronic kidney disease.

## Materials and Methods

### Kidney samples

Normal (non-cancer) Kidney tissue samples were obtained from surgical nephrectomies across multiple institutions via the Cooperative Human Tissue Network. The tissue was obtained from at least 2 cm from the tumor margin. An honest broker de-identified the samples and obtained clinical and demographic records from the medical records. The eGFR was recalculated using the CKD-epi 2021 equation^47^. The use of these samples and data was approved by the institutional review board of the University of Pennsylvania and deemed exempt as no identifiable information was collected.

### Kidney tissue RNA-seq and data processing

RNA isolation, sequencing and analysis were performed as previously published^29,48^. Total RNA was isolated from cortex kidney tissue using the RNeasy mini kit (Qiagen, Venlo, Netherlands) according to the manufacturer’s instructions, including the DNase digestion step. RNA quality was assessed by Agilent Bioanalyzer 2100. The cDNA library was prepared using NEBNext Ultra II RNA Library Prep Kit for Illumina. Then, cDNA libraries were sequenced on an Illumina NovaSeq 6000 platform using the NovaSeq PE150 protocol. Reads were aligned to the human genome (hg19) using STAR (v2.7.3a)^49^. Samples were selected that didn’t have evidence of autoimmune or cystic disease on pathology and had RIN >4.5, Mitochondrial RNA percent of <50, ribosomal RNA <10, Uniquely mapped percent more than 90% and sufficient total RNA count. Additionally, outliers were identified by PCA of mahalanobis distance and removed. 404 human kidney samples passed the above and were included in downstream analysis.

Raw counts of RNA expression were quantified using RSEM (v1.3.0). Counts were normalized using the Trimmed Mean of M-values (TMM) method implemented in the edgeR^50^ package, and a voom transformation was applied to obtain log2 counts per million (log-CPM), weighted to account for differences in variance across expression levels. Genes were filtered to retain those with at least 25% of values exceeding a value of 0.1. Quantile normalization was then performed. Transcript expression was obtained from the isoform quantification by RSEM(v1.3.0) and processed similar to raw counts.

To obtain quantification of splicing events, we used LeafCutter (v0.2.9)^15^ that uses junction spanning reads to infer spliced introns. After alignment by STAR, Junctions were annotated from the bam files using the junction extract function of Regtools(v1.0.0)^51^. The resulting junction files were then used by leafcutter to quantify IER. Clusters were selected that had at least 50 aligned reads and a maximum intron length of 500 kilobases. The resulting fractions were converted to proportions of each intron isoform. Clusters were filtered if present in less than half of the samples and if there was low variation (abs Z-score, less 0.25 in n-3 samples or abs Z score >6 in less than 3 samples)^28^.

To identify the proportion of the types of alternative splicing events, we used rMATs turbo^52^. This quantified the events into mutually exclusive exons, skipped exons, intron retention and alternative 5 and 3 prime splice sites. The number of significant events was counted in each output file and displayed as a percentage of the total. To assess the association of splicing with disease, the count file of the intron excision ratio of each intron isoform was transformed using arcsine transformation of the square root of the value. And a linear regression was used for estimated glomerular filtration rate (eGFR) as an outcome adjusting for age and sex as covariates. Similarly, log transformed interstitial fibrosis (incremented by 1%) was also used as an outcome for 225 samples that have interstitial fibrosis scored. The p value was adjusted using false discovery rate (FDR). The beta coefficient was then back transformed to identify the beta coefficient of the outcome to the change in intron excision ratio from 0.5-0.6. Visualization of the sashimi plot was performed by ggsashimi^53^ Sex specific differential splicing was performed using leafcutter_ds.py on the count matrix that was filtered based on above.

### Kidney Tissue Proteomics and data processing

Proteomics dataset used in this study was recently published. Briefly, for pQTL the Somascan platform was on human kidney cortex tissue from 325 samples resulting in the quantification of 6,903 proteins. The majority of the samples are also part of the same cohort used for RNA sequencing (281 samples). Inverse transformed quantile normalized expression matrix was used for cis-QTL mapping with whole genome sequencing of these samples using QTLTools. The results of this were used to prioritize genes with a shared regulatory effect on splicing, RNA expression and protein expression. For protein abundance, liquid chromatography mass spectrometry (LC-MS) was performed using the Orbitrap Eclipse mass spectrometer coupled with FAIMS and Real-Time-Search using the protocol detailed in a recent publication^54^.

### Whole genome sequencing and variant calling

Detailed description of the whole genome sequencing and variant calling of kidney tissue was previously published. Briefly, DNA was extracted from kidney tissue using the DNeasy blood and tissue Qiagen kit (QIAGEN) according to the manufacturer’s instructions and aligned to hg19 reference genome using Burrows-Wheeler Aligner (BWA) v0.7.17^55^. After quality control, GATK haplotype caller was used for variant calling and variant quality score recalibration was performed using GATK best practices^56^.

### Genotype data quality control

SNPs were filtered out if missingness was higher than 5%, they had a MAF less than 5%, or if the significance level for the Hardy-Weinberg equilibrium (HWE) was less than 1 x 10^-6^. In addition, samples were removed in the case of across-SNP call rate less than 95%, sex mismatch and heterozygosity if Z score is 3 or more. Relatedness was determined by inferring Inheritance by descent using KING v2.3.1 ^57^ and samples were up to 2^nd^ degree relatedness were removed.

### Molecular cis Quantitative Trait Loci

#### Expression QTL (eQTL)

Quantile normalized files were inverse normal transformed. Age, biological sex, batch, site and Total RNA read and RNA integrity number (RIN) were used as covariates. To account for population structure, the first 6 genotype principal components were used. PEER factors were used to account for hidden covariates^58^. The number of PEER factors was chosen that maximizes the number of significant QTLs. 45 peer factors were used for short read RNA count expression. TensorQTL^59^ was used for QTL analysis using 1Mb window. First, a permutation run was performed to identify the most significant variant for each gene. Storey’s qvalue^60^ was calculated and Genes with a qvalue less than 0.05 were considered eQTL genes. Nominal p values were calculated by running a cis_nominal function in TensorQTL and SNPs were considered significant if the nominal p value was less than the gene level q value. Transcript expression QTL (teQTL) was performed in a similar method to eQTL but the phenotype was grouped by gene using the -- phenotype-groups function in tensorQTL. A gene was considered teQTL gene if any transcript had a qvalue less than 0.05. Splicing QTL (sQTL) was performed in a similar way. The quantile normalized count matrix was inverse normal transformed. The phenotype was grouped by intron clusters and a gene was considered an sQTL gene if any intron cluster had a qvalue less than 0.05. SNPs were considered significant if they had a nominal p value that was less than the cluster level q value. Variant annotation and prediction of the functional SNP effect of significant eSNPs and sSNPs was performed using SnpEff v5.2c^61^. Variants in the 95% credible sets for each significant QTL was obtained using susieR package and the annotations were compared between sQTL and eQTL. ATAC seq results were obtained from bulk assay for transposase-accessible chromatin (ATAC)–based allele-specific accessibility^22^ with chromHMM annotations from kidney chromatin data obtained from encode^62^. The QTL SNP variants were intersected to identify if they lie in open peaks and the functional annotation.

### Colocalization

Bayesian colocalization was performed using an eGFR GWAS^22^. Results of 1,775,389 subjects of European ancestry were used. Coloc v5.2.3^63^ R package was used using default parameters of coloc.abf of a prior probability of 1e-4 of association with either trait and 1e-5 of association with both traits. 100kb genetic region was used on either side of the sentinel SNPs in each locus. Summary statistics of overlapping SNPs (Beta SNP effect and standard error) in eGFR GWAS and each of eQTL and sQTL. Only regions with ≥ 30 SNPs were included in the analysis. A posterior probability of a SNP being associated with both traits (PP H4) of ≥ 0.8 was used as a cutoff. The prior probability was adjusted for loci that met this threshold and the posterior probability recalculated. For sQTL, genes with any intron isoform that exceeds this cutoff is considered a coloc gene.

Multitrait colocalization was performed using moloc v0.1.0 R package^64^. The default prior variances of 0.01, 0.1, 0.5 and prior probability of trait associations of 1e-04,1e-06 and 1e-07 were used. GWAS, sQTL and eQTL were used as traits. Regions were defined similar to coloc but regions with ≥ 10 SNPs were used for the analysis and a cutoff of PPH4 of 0.8 was used as well. pQTL results from a previous study were used with GWAS and SQTL as well. Overlapping genes between these two multitrait colocalization were identified.

### Summary Mendelian Randomization (SMR)

To further prioritize QTL genes for kidney function, we used summary mendelian randomization analysis on the eGFR GWAS and the eQTL/sQTL using the SMR command line tool^65^. Significant SNPs in sQTL genes/eQTL genes were used for each QTL dataset and converted to binary format (BESD). Variants in chromosome 6 that correspond to the HLA locus were excluded to reduce potential confounding. The European population of the 1000 genomes project^66^ was used as a reference for LD estimation as 88% of the study cohort were of European ancestry. To account for heterogeneity of instrument effects, Heterogeneity in Dependent Instruments (HEIDI) test was performed using a p value cutoff of 0.01. Bonferroni correction was applied to the SMR p value to account for multiple testing by dividing 0.05 by the number of testing genes.

### Cardiac Kidney and Metabolism GWAS traits

To identify if Intron isoforms in the kidney are related to extrarenal cardiac and metabolic traits, we utilized publicly available GWAS summary statistics for Coronary Artery Disease^67^,Heart Failure^68^, Systolic and Diastolic blood pressure^69^, Lipids to include HDL, LDL and Triglycerides^70^, obesity^71^, Type 1^72^ and Type 2^73^ diabetes mellitus. SMR command line tool and COLOC were used in a similar manner as above to prioritize causal sQTL genes with an effect on each outcome. To define the loci, a window of 200kb was created from the transcript start site of each sQTL gene using genomic ranges ^74^ and colocalization was performed if a variant had a p-value less than 5e-10. The results were compared with the eGFR sQTL genes. For the MANBA and ERAP2 gene, the SNPs for the MANBA probe was run using the – extract-exposure-probe option.

### Single Cell Sequencing

To identify if sQTL genes are localized to certain cells in the kidney, we utilized a single nuclei sequencing dataset in human kidney tissue^75^(accessed from https://www.ncbi.nlm.nih.gov/geo/query/acc.cgi?acc=GSE211785). The data was reanalyzed with consolidation of the clusters into 9. The mean expression of each gene was obtained using the Scanpy package^76^ and normalized across each gene. The results were then plotted on a heatmap to compare the relative expression in each cell type. snATACseq was obtained from previously published data of 6 adult human kidney samples in 57,229 cells^48^. (accessed from https://susztaklab.com/index.php).

### Long Read RNA Sequencing

Long read sequencing was performed on 8 kidney cortex samples, 4 with eGFR < 60 and 4 with eGFR >60. RNA was extracted as above. The Pacbio Kinnex full-length RNA kit protocol was used for poly-A primed full length cDNA generation and HiFi sequencing on two Revio Single Molecule, Real-Time (SMRT) cells per manufacturer’s instructions. The sequenced files were analyzed according to the Pacbio Isoseq pipeline^77^. Skera was used to deconcatenate the reads from each SMRT cell HiFi bam file. Primers were removed from the segmented bam files and demultiplexed using lima. Isoseq -refine was used to trim poly A tails and remove concatemers from each sample bam files to generate full length non-concatemer (flnc) bam files. Isoseq cluster2 was used to cluster the full length (FL) reads and only includes isoforms with at least 2 FL reads. The clustered reads were aligned to the hg38 reference genome using pbmm2. Similar isoforms in the clustered reads were collapsed using isoseq collapse. Isoforms were filtered using Pigeon to remove transcripts that are likely artefact by intrapriming, mono-exonic and having low coverage of less than 3 per junction. Pigeon Classification was used to classify the isoforms using the SQANTI classification^78^. Full Splice Match (FSM) isoforms match fully to reference isoforms whereas Incomplete Splice Match (ISM) partially align to the reference. Novel isoforms were classified as Novel in Catalog (NIC) contain new combinations of already annotated splice junctions or novel splice junctions formed from already annotated donors and acceptors, whereas Novel Not in Catalog (NNC) use novel donors or acceptors. Additional categories include intergenic (outside boundaries of an annotated gene), genic genomic (partial intron/exon overlap in a known gene) and Fusion (transcript spans two annotated loci). The collapsed fasta files were demultiplexed and only isoforms present in 3 or more samples were included. Plot of isoforms was performed using ggtranscript v1.0.0 R package^79^. Introns were shortened for visual clarity. To compare the CKD and control group, tappAS v 1.0.8^80^ was used. Isoforms not expressed in all samples were filtered. DEXSeq^81^ was used for differential isoform usage and NOISeq^82^ for differential gene expression within the tappAS platform. Enrichment was performed using DAVID^83^.

## Data Availability

Raw data, processed data, and metadata from the single nuclear RNA-seq and spatial RNA-seq have been deposited in Gene Expression Omnibus (GEO) with the accession code of GSE211785. The single nuclear expression and spatial data is also available at www.susztaklab.com (https://susztaklab.com/hk_genemap/). No consent was obtained to share individual-level genotype data.

## Author Contributions

S.M. analyzed the data. D.H., E.H., A.A., A.B. and B.D. assisted with data generation and analysis. K.S. was responsible for overall design and oversight of the analysis. D.H. generated the pQTL data. F.E.M. and T.N. generated the LC-MS data. S.M. and K.S. wrote the original draft. All authors approved the final version of the manuscript.

## Acknowledgement

We thank Glennis Logsdon, PhD (Department of Genetics, University of Pennsylvania) for helpful input in long read sequencing.

## Code availability

All codes used for the analysis are provided in a Github repository https://github.com/samohandes/splicing_ckd. Table 1.

---

**Extended Data 1.**
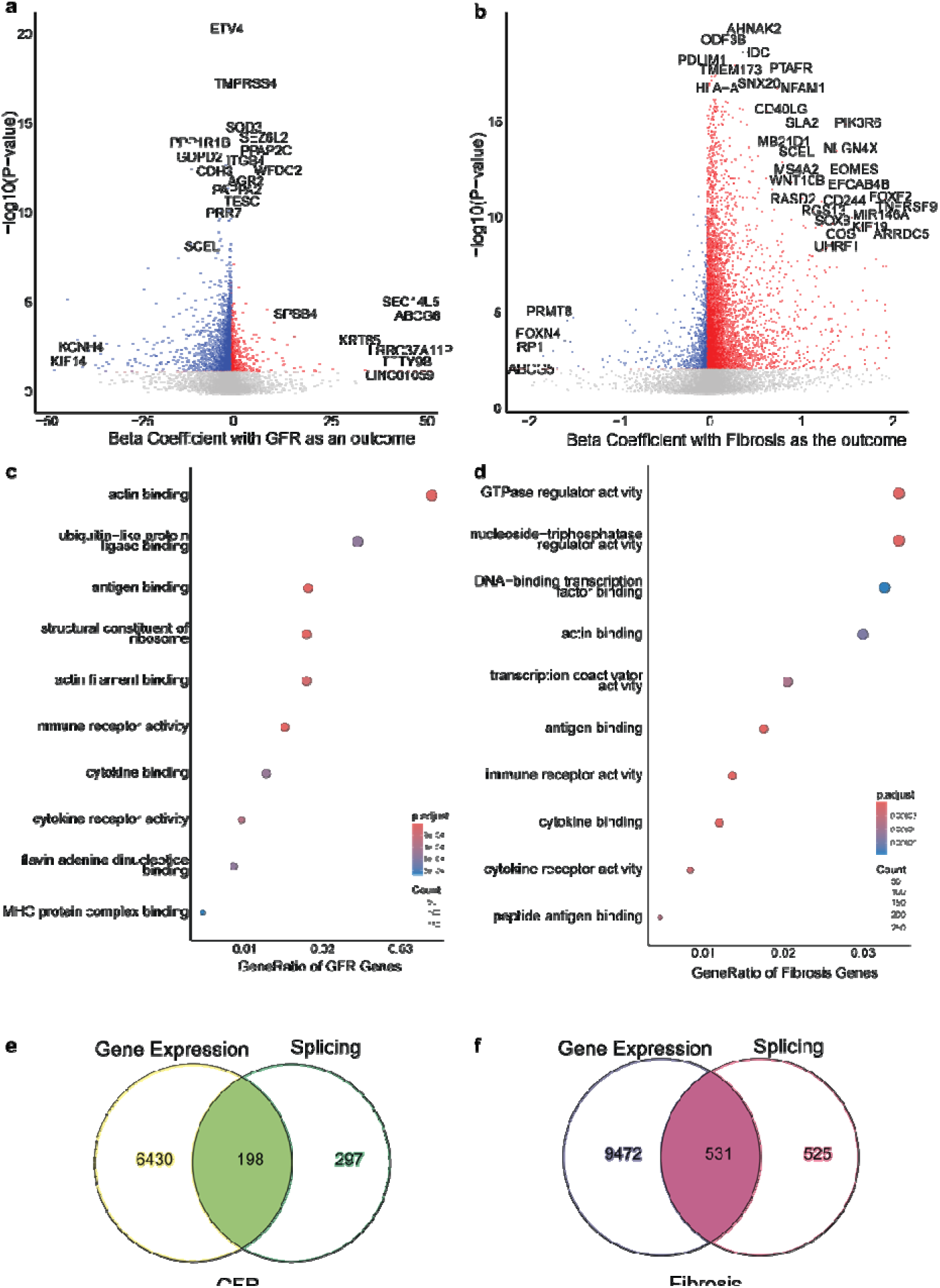
Association of RNA expression (TPM) with kidney disease. A. Volcano plot of the beta coefficient of each gene in a linear model with GFR and B. with log Fibrosis using Age and Sex as covariates. Each dot represents a gene and genes significant at a p-value of 0.05 after FDR correction are colored. Dot plot of the functional pathways enriched by significant genes associated with C. GFR and D. Fibrosis. Comparison of the overlap of the significant genes by RNA expression and Splicing with E. GFR and F. log Fibrosis showing around half the genes that have a significant splicing association also have a significant change in total RNA abundance.

**Extended Data Figure 2.**
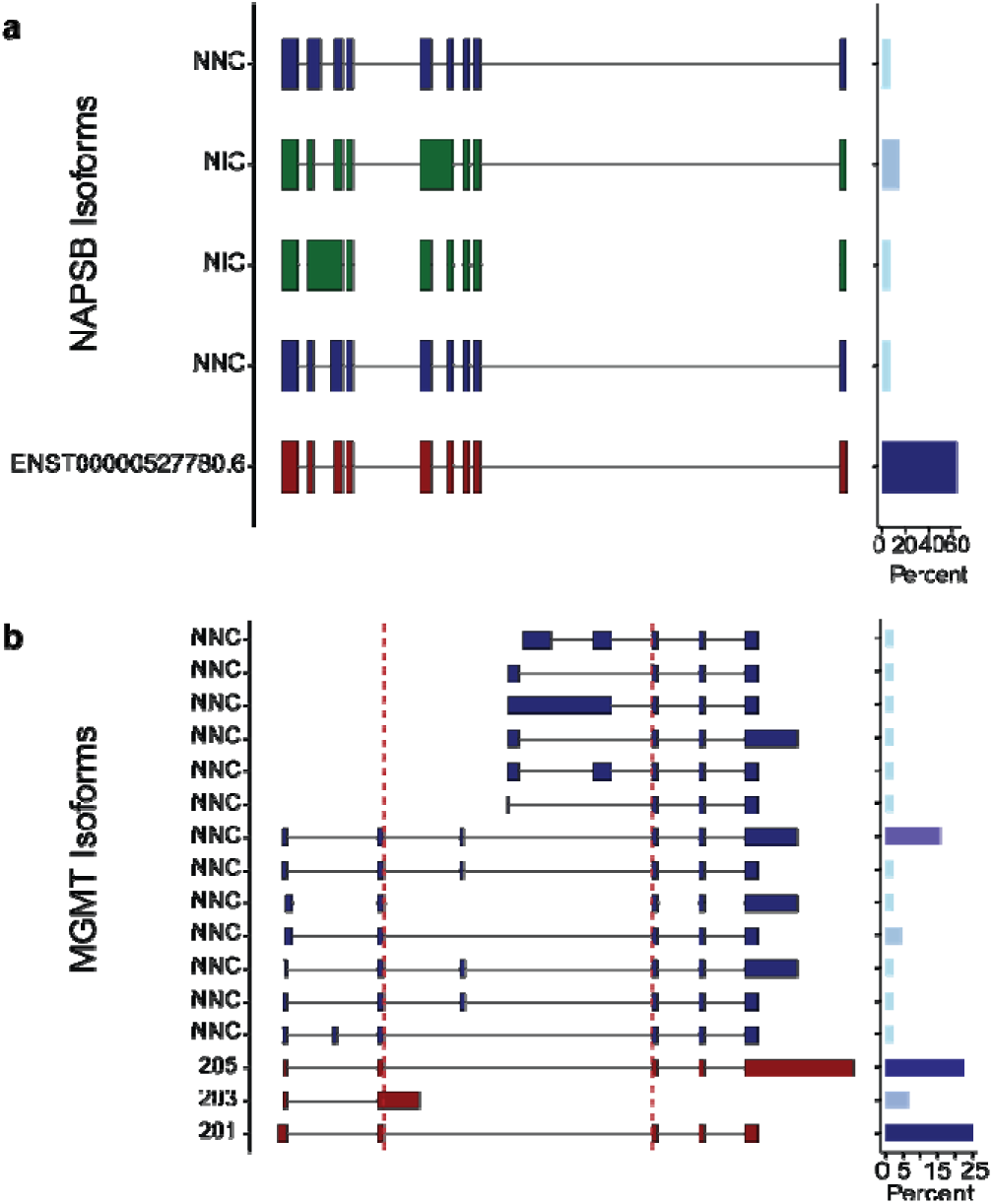
Long read isoforms of A. NAPSB showing 1 known isoform and 4 novel isoforms. The known isoform accounts for approximately 60% of the transcript abundance. X-axis is the position on chromosome 19. Plot on the right shows the relative abundance of each isoform in long read sequencing. B. MGMT showing 13 novel isoforms and 3 known isoforms. Dashed line in red shows the significant junction by sQTL.

**Extended Data Figure 3.**
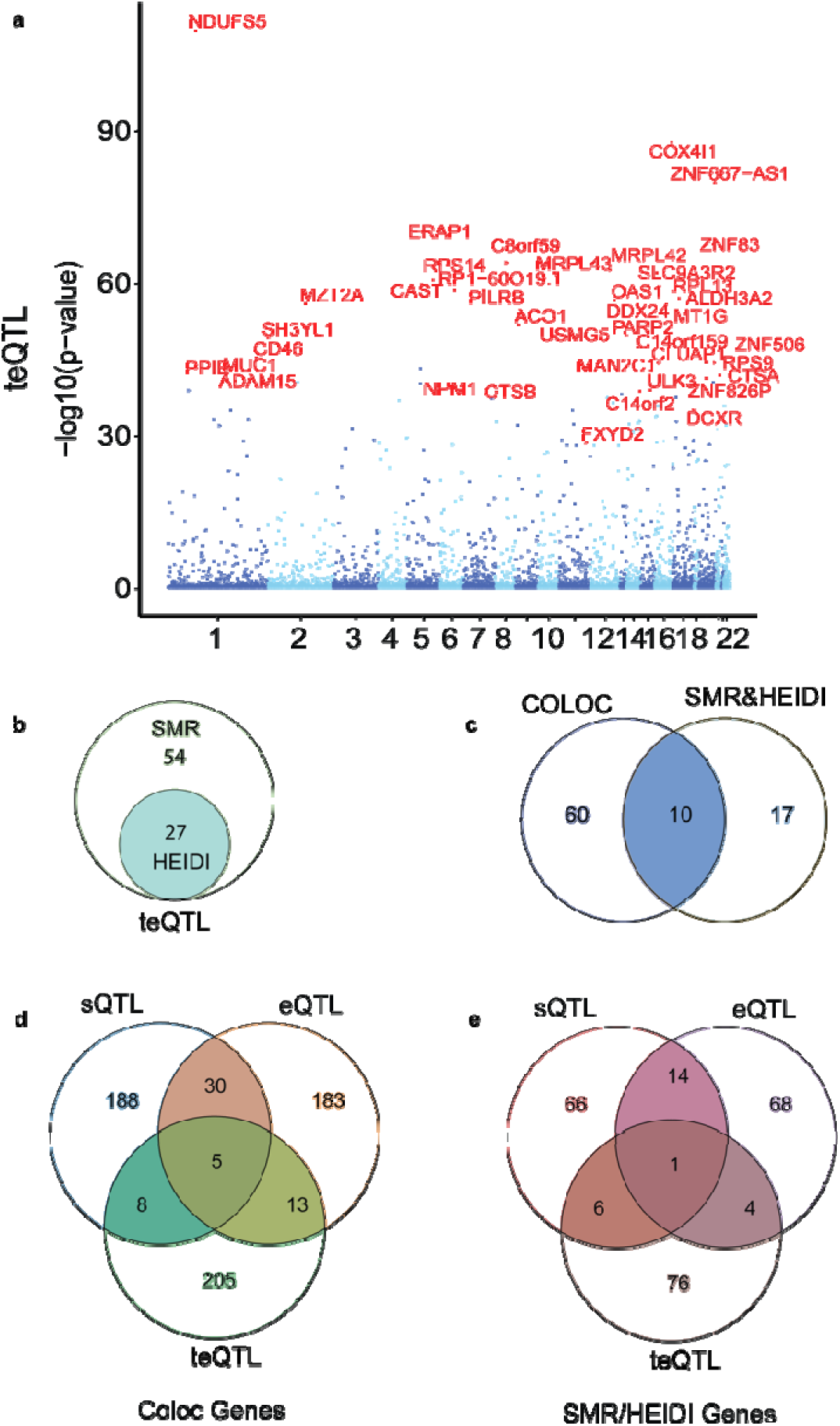
A. Manhattan plot of teQTL genes. Each dot represents a genetic variant. x-axis is the location on the reference genome grouped by chromosome. Y-axis is -log10 p value. B. SMR and HEIDI of teQTL genes nominating 27 genes. C. Overlap of COLOC and SMR genes in teQTL nominating 10 genes significant by both methods. Overlap in sQTL, eQTL and teQTL genes from D. COLOC and E. SMR/HEIDI genes. minimal overlap is seen between teQTL and sQTL datasets.

**Extended Data Figure 4.**
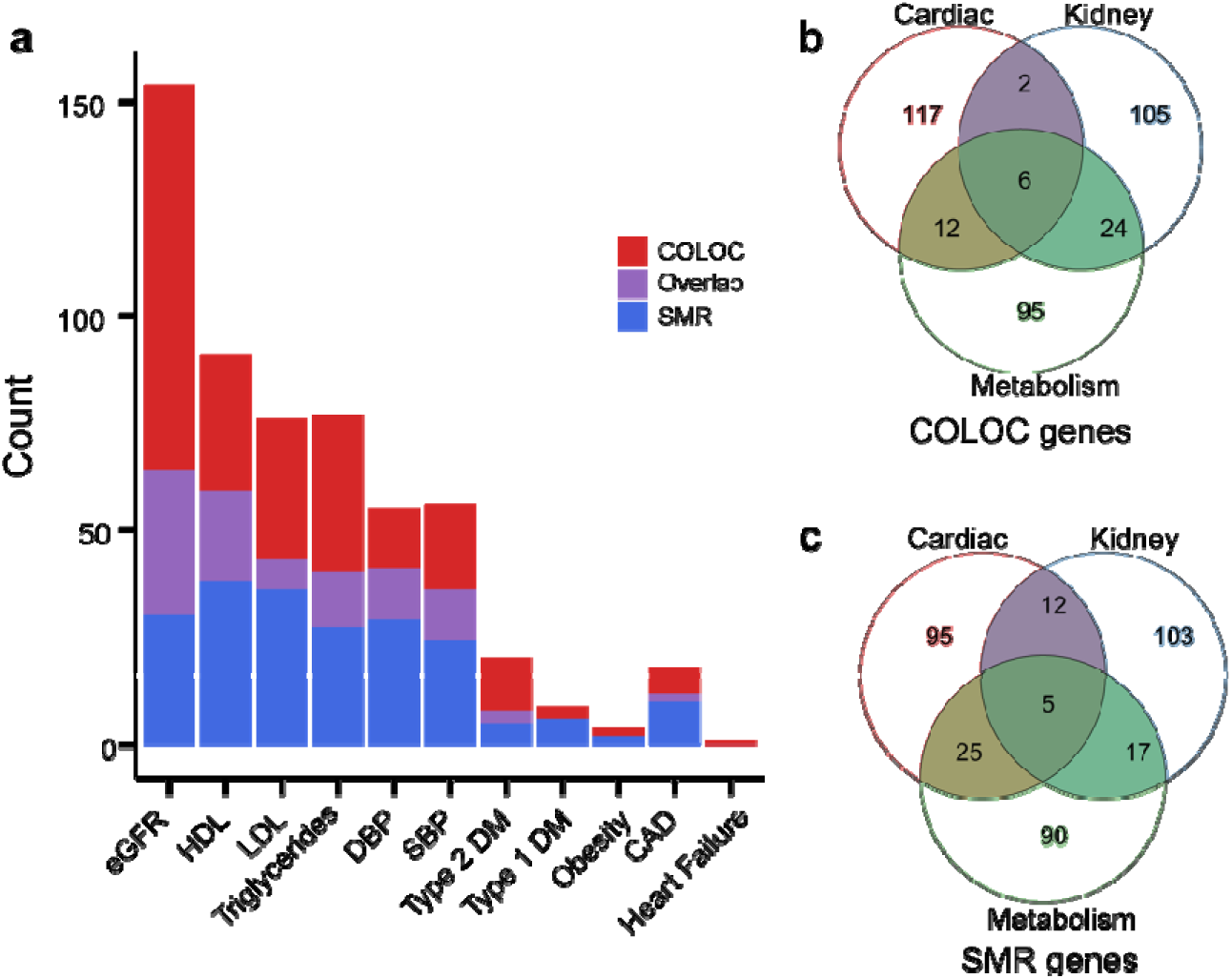
A. SMR and Coloc of CKM syndrome traits. Genes nominated by SMR (blue), COLOC (red) and the overlap (purple) are shown. Overlap of B. COLOC genes and C. SMR genes in Cardiac, Kidney and Metabolic syndrome traits. These demonstrate shared gene regulation in the kidney between among traits in the syndrome highlighting the shared pathophysiology.

**Extended Data Figure 5.**
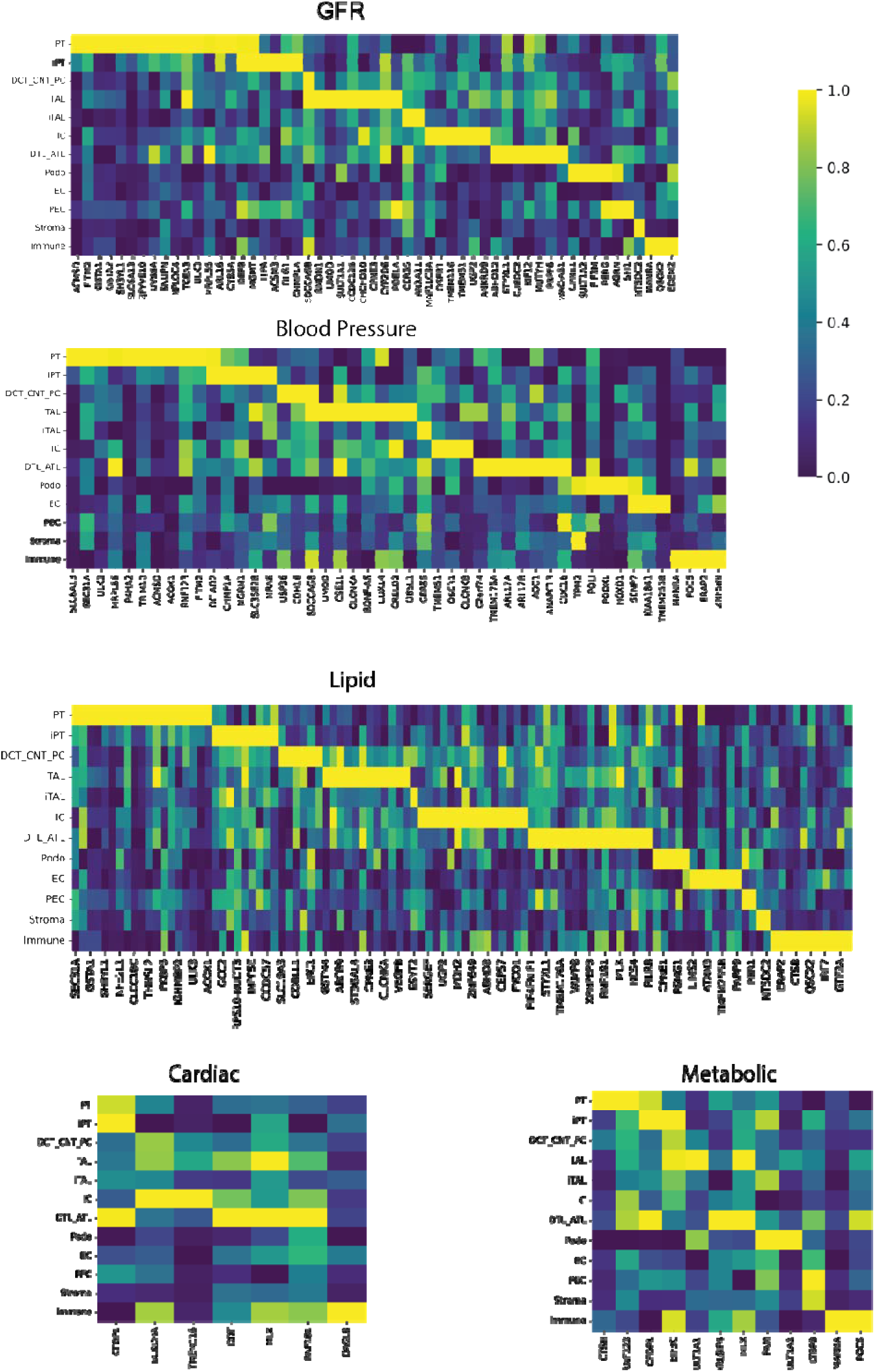
Single Nucleus RNA expression of sQTL genes in CKM traits. nominated by A. GFR B. Blood Pressure C. Lipids D. Cardiac and E. Metabolic traits. X-axis is the genes and the y axis is the unique cells in the single cell dataset. The expression is scaled by each gene to show the relative expression of each gene by cell type with the highest in yellow and the lowest in dark blue.

