## Supplementary Figures for "Human Kidney Alternative Splicing Information Illuminates Cardiovascular-Kidney-Metabolic Syndrome Risk"

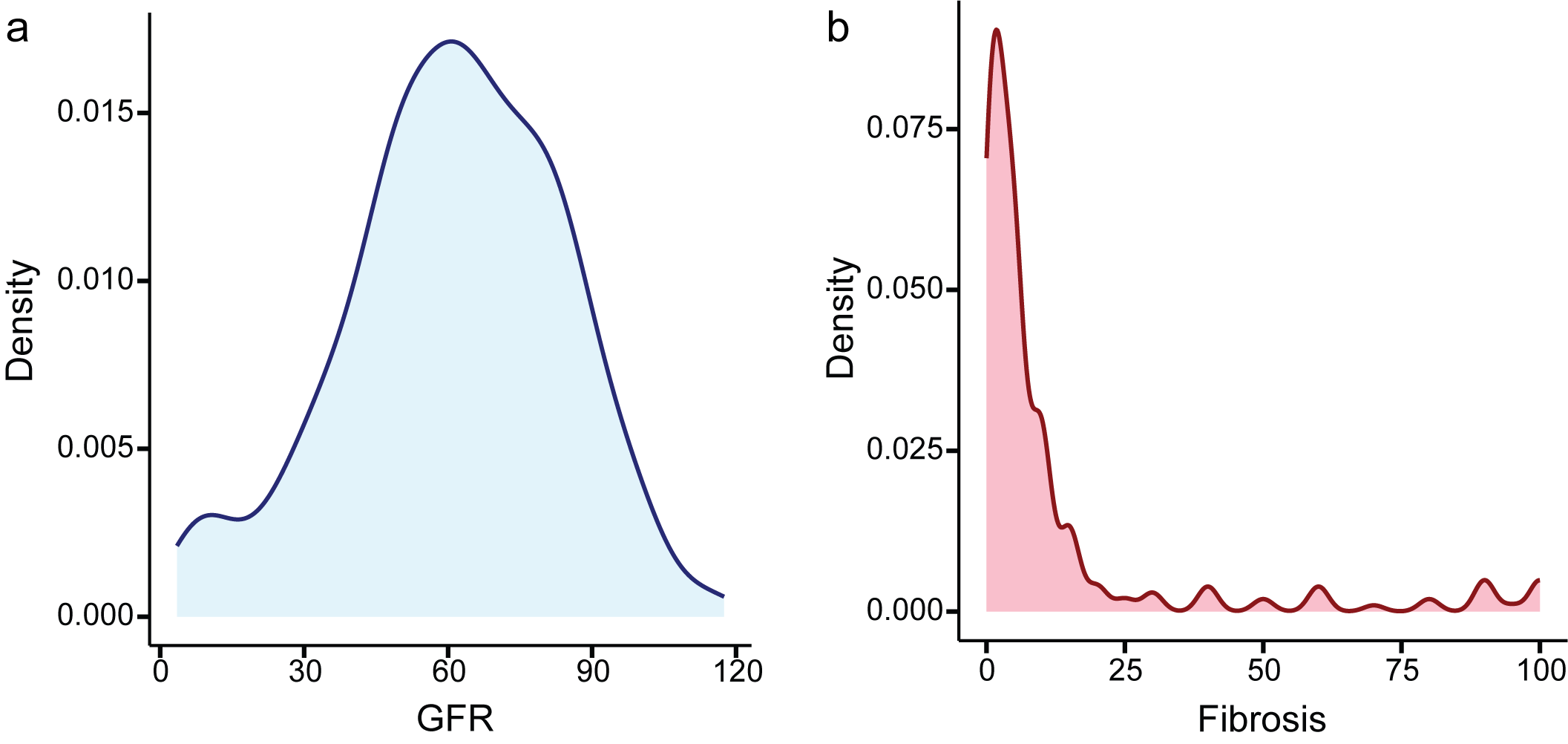

Supplementary Figure 1 A. Density plot of GFR showing an apparent normal distribution. B. Interstitial fibrosis, however, is significantly skewed to the right. The x-axis shows the GFR and Fibrosis values respectively whereas the y-axis shows the density.

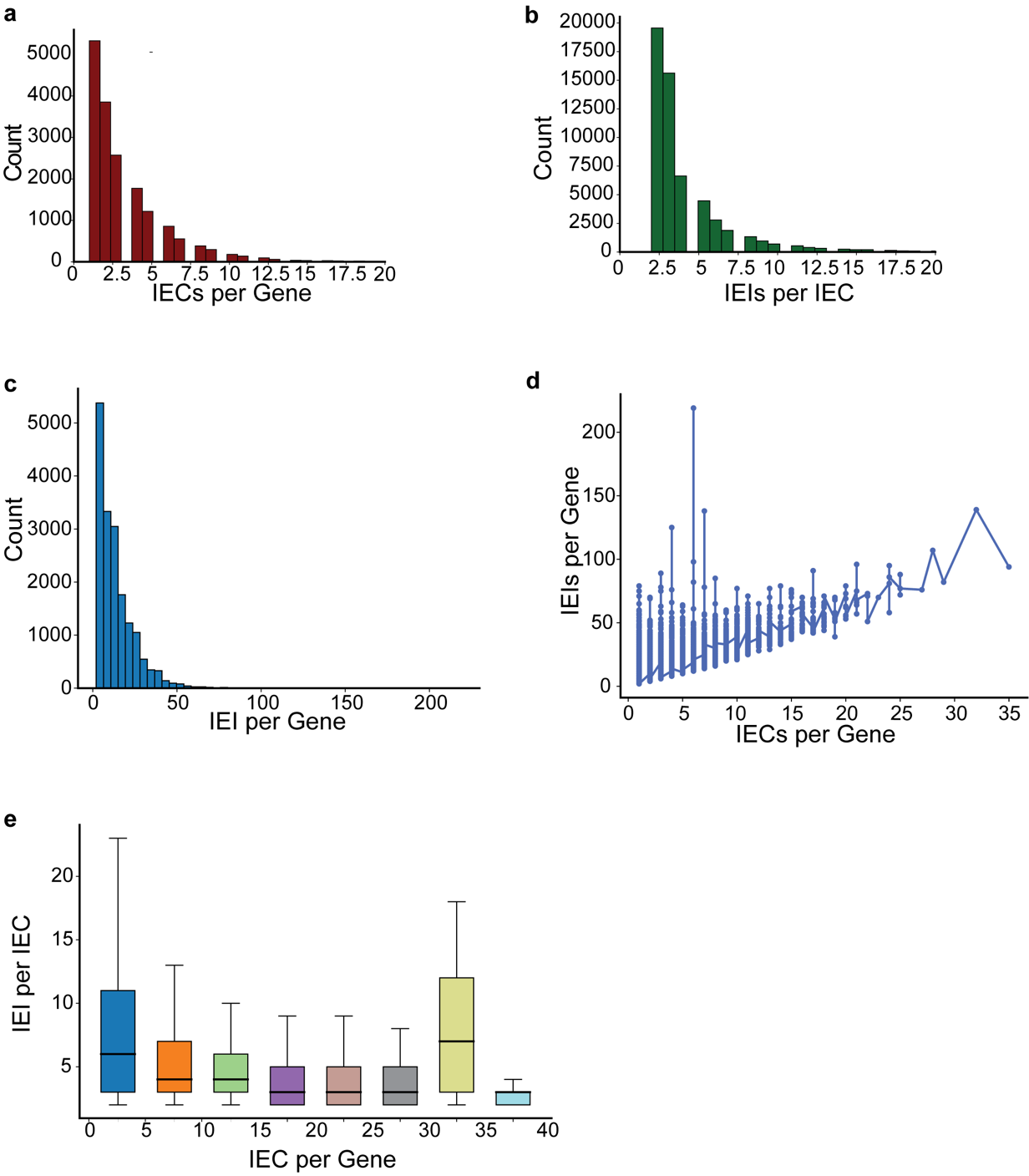

Supplementary Figure 2. A. Distribution of the number of intron excision clusters per gene (x-axis). The median is 3 and the distribution is skewed to the right. B. Count of the distribution of the number of intron excision isoforms per intron cluster (x-axis). The median is 3 and the distribution is skewed to the right. C. Distribution of the number of intron excision isoforms per gene (x-axis). The mode is 1 intron isoform per gene. D. intron clusters per gene (x-axis) compared to intron isoforms per gene (y-axis). This shows a linear increase in number of intron isoforms as the number of intron clusters increases. E. intron clusters per gene (x-axis) compared to intron isoforms per cluster (y-axis).

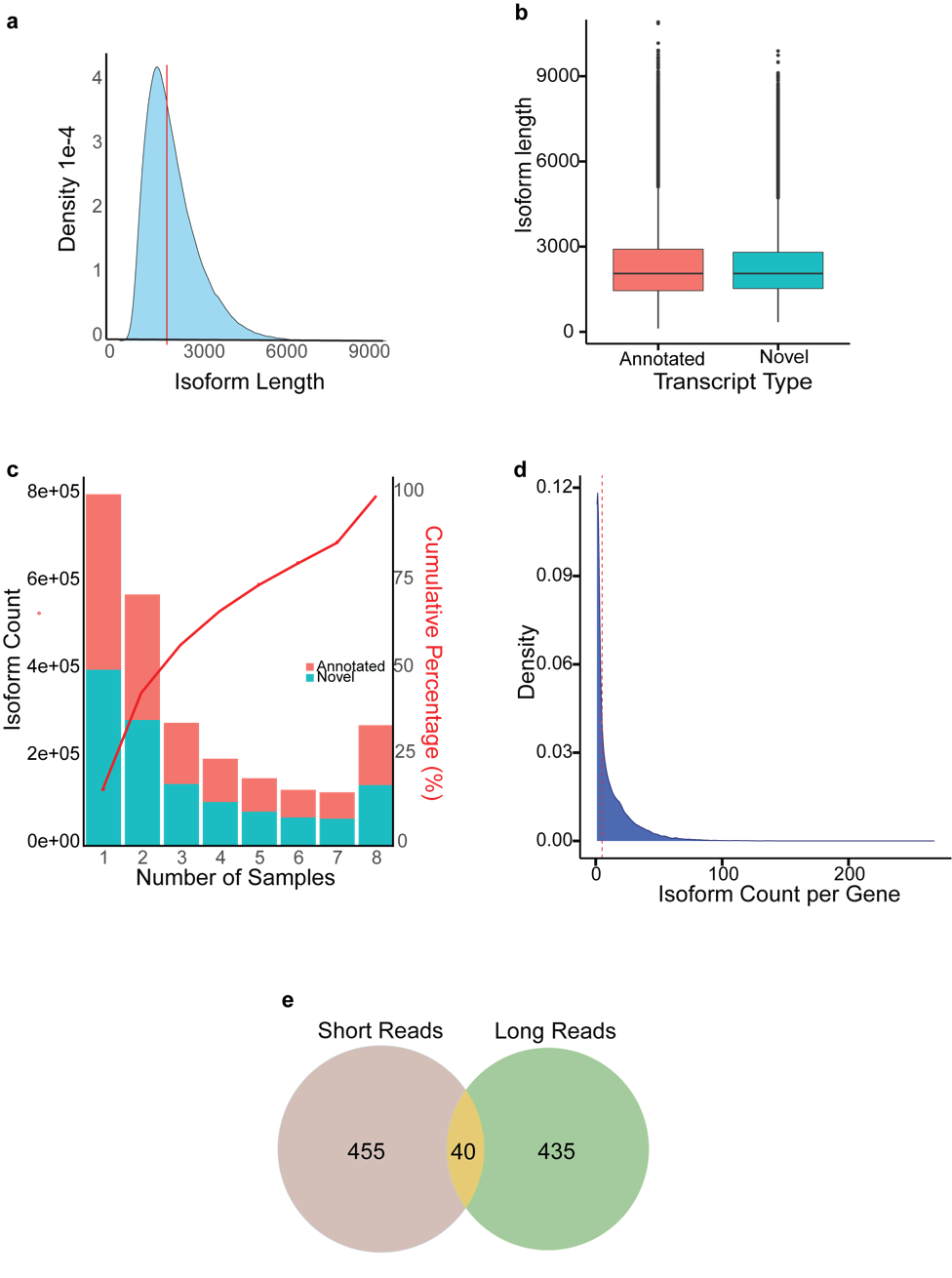

Supplementary Figure 3. A. Density plot of the length of the long read isoforms. X-axis isoform length, y-axis density. B. Difference in length between annotated and novel isoforms. No significant difference in gene length was seen between these two groups. C. Distribution of number of samples represented by each isoform. X-axis is the number of samples expressing each transcript. Each bar is grouped by annotated isoforms (red) or novel (blue). A large proportion of isoforms are present in 1-2 samples only. Right y axis shows the cumulative percentage of the transcripts expression distribution by number of samples. This shows an apparent inflection point at 2 samples.D. Density plot of the number of isoforms per gene. Isoforms present in less than 3 samples were excluded. The median count is 5 and highlighted in red. E. Overlap of significant differential isoform usage between long reads and short reads. A small portion are overlapped using these different approaches.

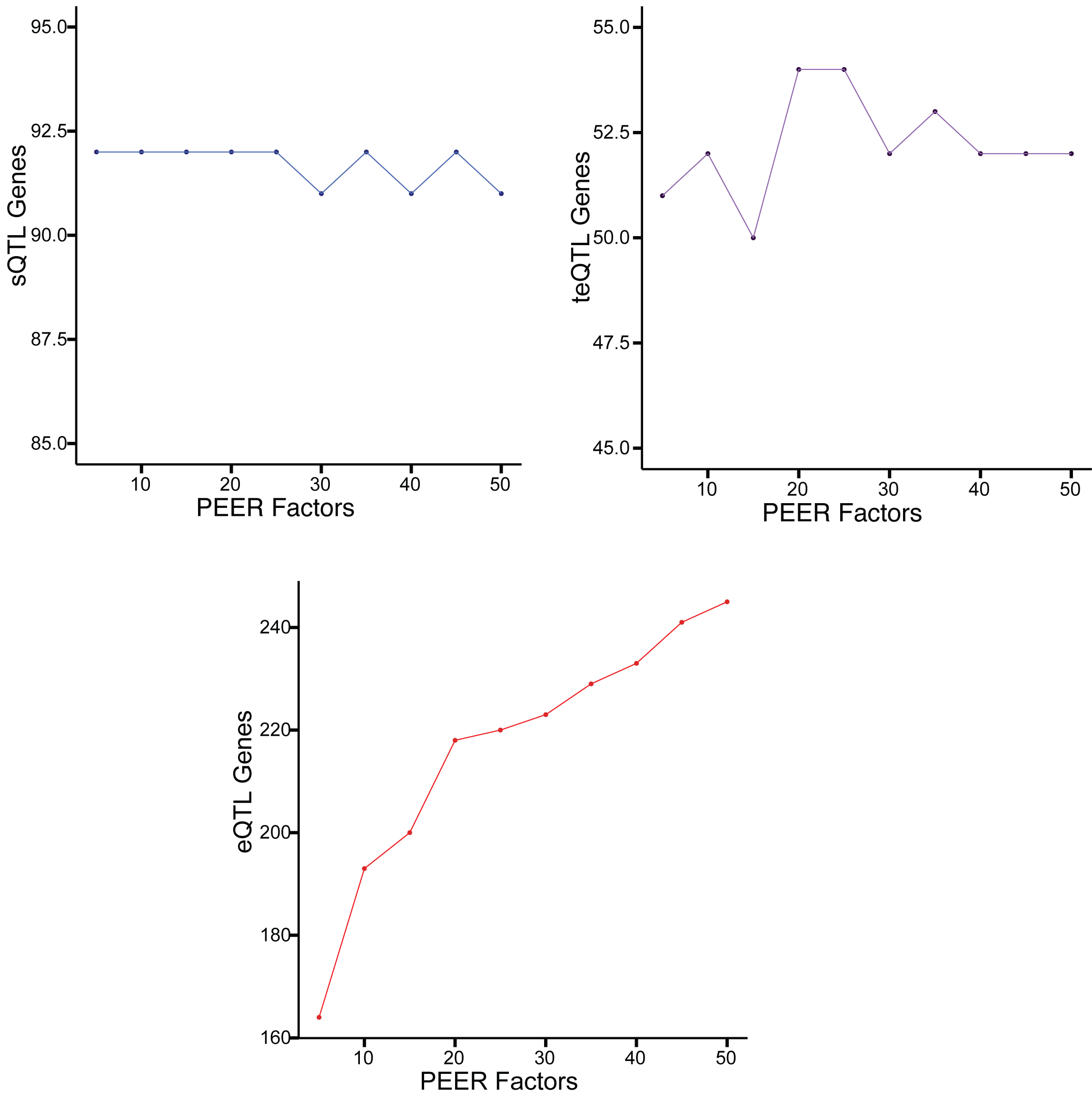

Supplementary Figure 4. Significant Genes in Chromosome 10 by peer factors in sQTL, teQTL and eQTL.x-axis are the PEER factors and y -axis show the number of significant respective QTL genes. eQTL genes showed a significant change by PEER factors but sQTL and teQTL show limited change.

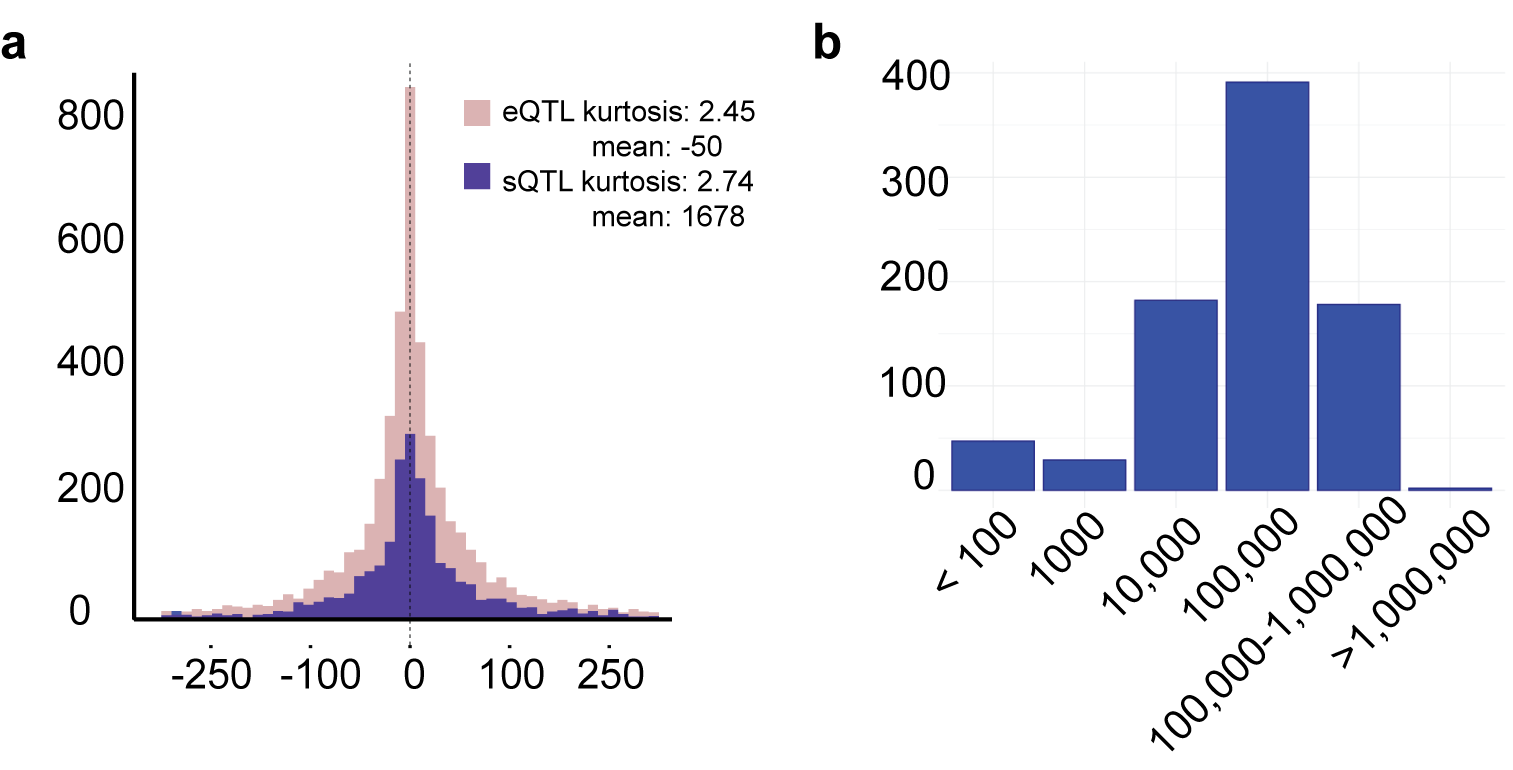

Supplementary Figure 5. A. Distance of best QTL variant to transcript start site from the tensorQTL permuation pass. The best variant is the one with the least p-value. X-axis is the distance in base pairs and the y axis is the number of variants. The mean distance of the sQTL variant is further from the TSS than eQTL. B. Distance between the most significant eQTL and sQTL variant in overlapping genes. x-axis is the distance in base pairs grouped into the shown distances due to the wide range. Y axis is the number of genes. The mode was around 100,000 base pair distance.

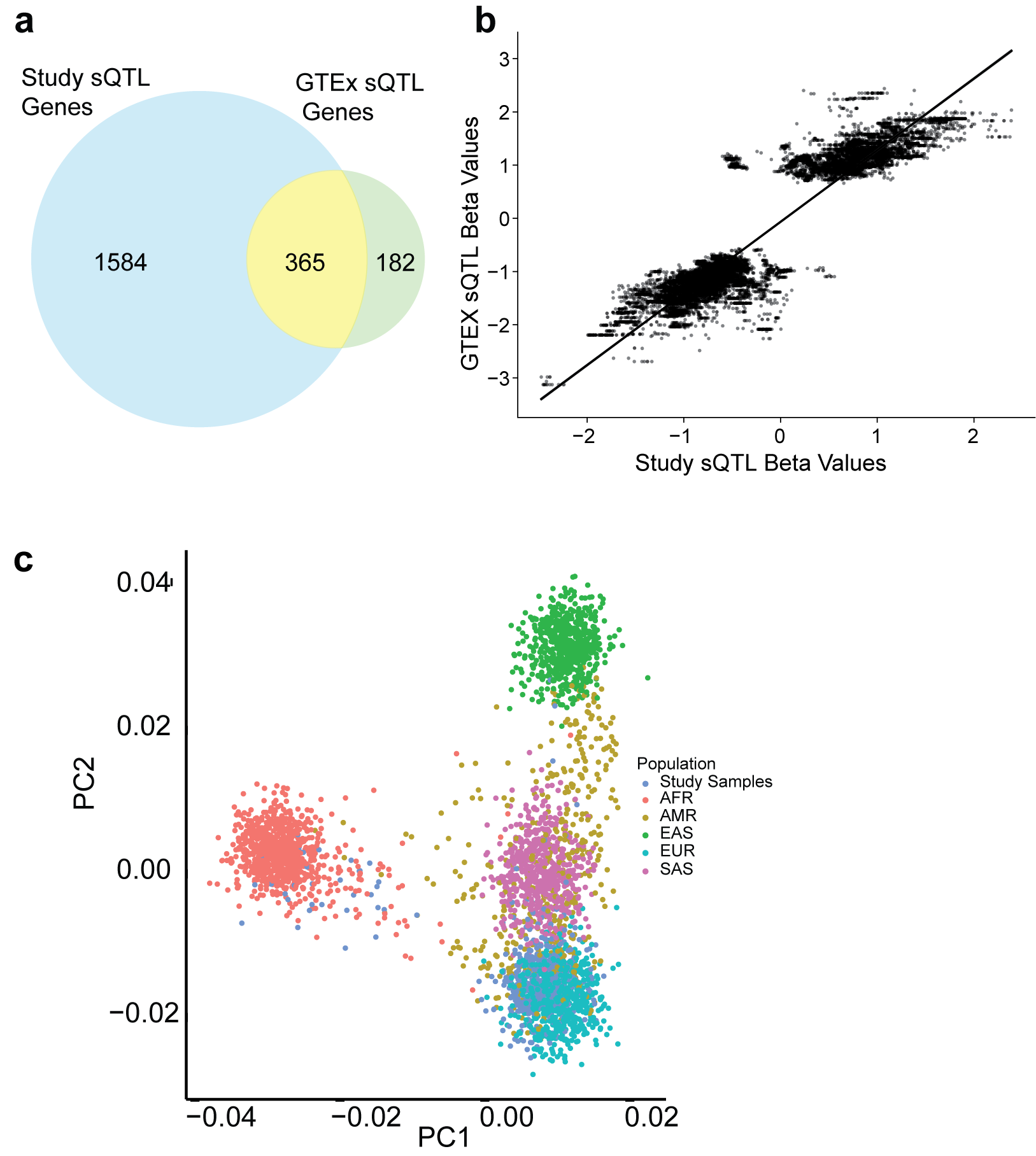

Supplementary Figure 6. Validation of sQTL results with GTEx v8 dataset. A. Overlap of significant genes, the majority of the genes found in the GTEx dataset were found in this dataset. B. Plot of beta coefficient of overlapping genes showed a good correlation. Each dot represents a gene variant. X-axis is the beta value of the study cohort and y axis the beta value of the GTEx cohort. C. PCA plot of the merged genotyping with the 1000 genome project. Each dot is a sample, the study cohort samples are in dark blue and the 1000 genome project samples are colored by the superpopulation designation. The majority of the study cohort grouped with the 1kgp EUR superpopulation, consistent with reported background.

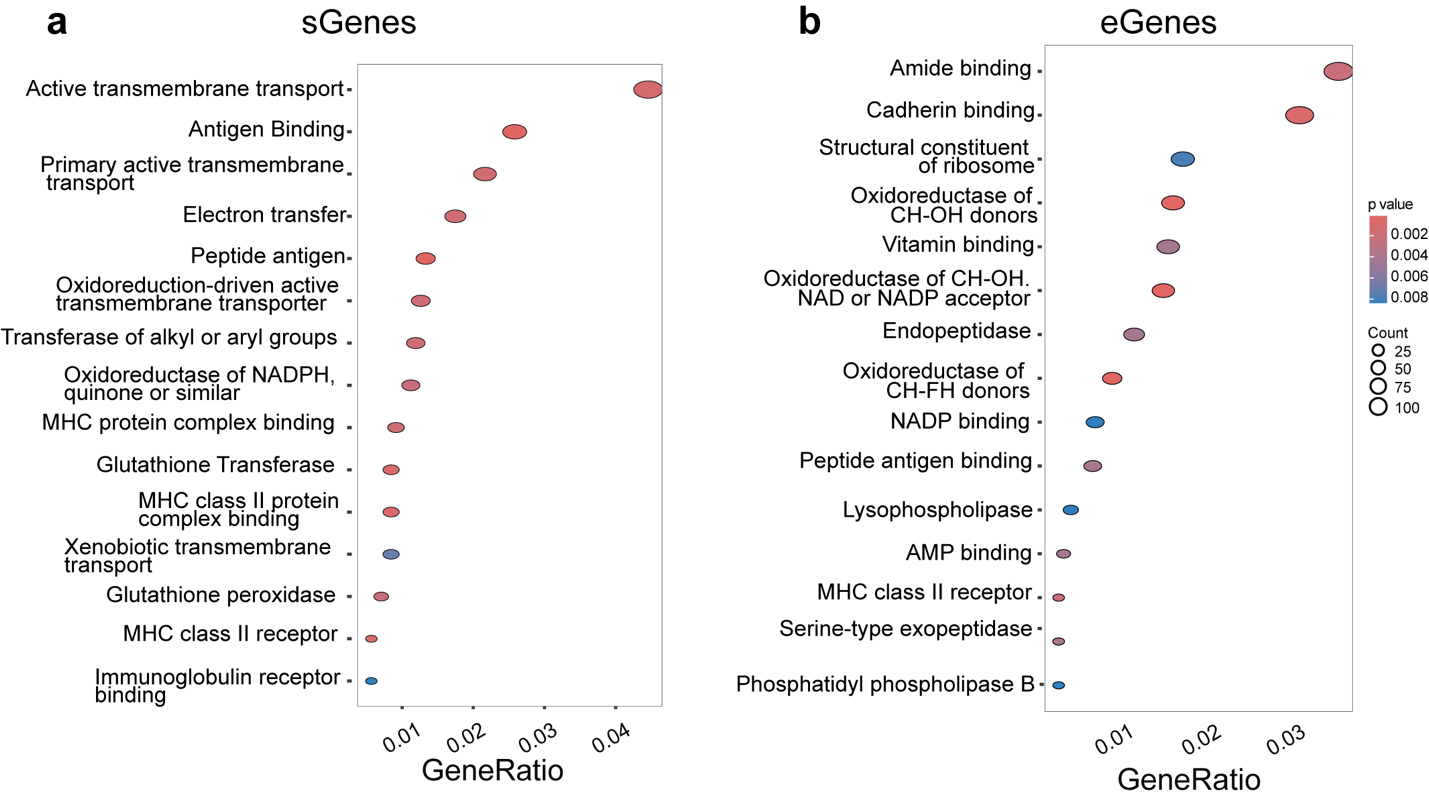

Supplementary Figure 7. Functional pathway enrichment of significant QTL genes in A. sQTL and B. eQTL

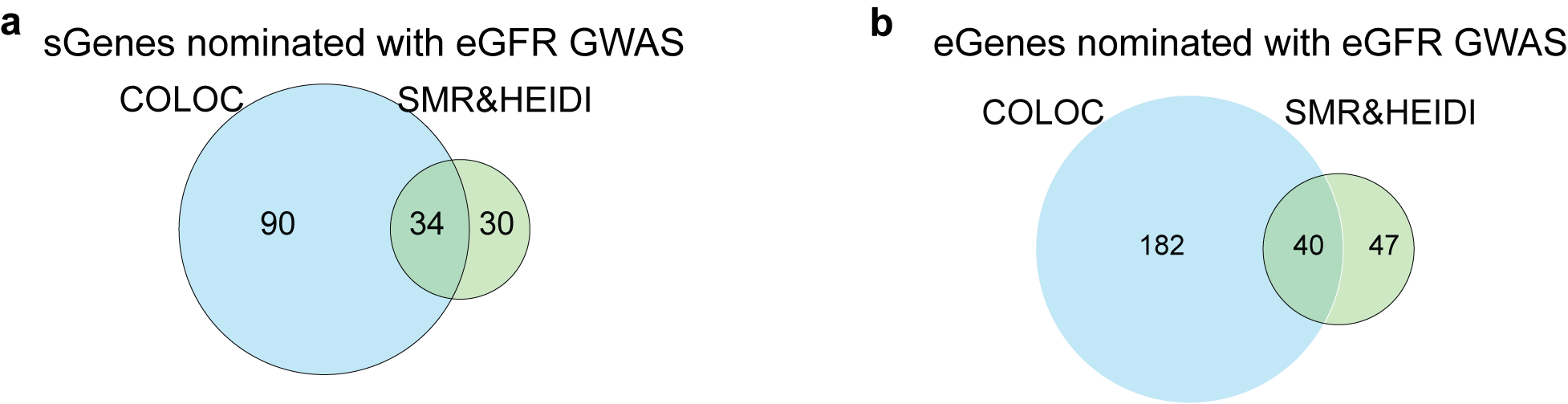

Supplementary Figure 8. Overlap of genes nominated by Colocalization and SMR&HEIDI in A. sQTL genes and B. eQTL genes

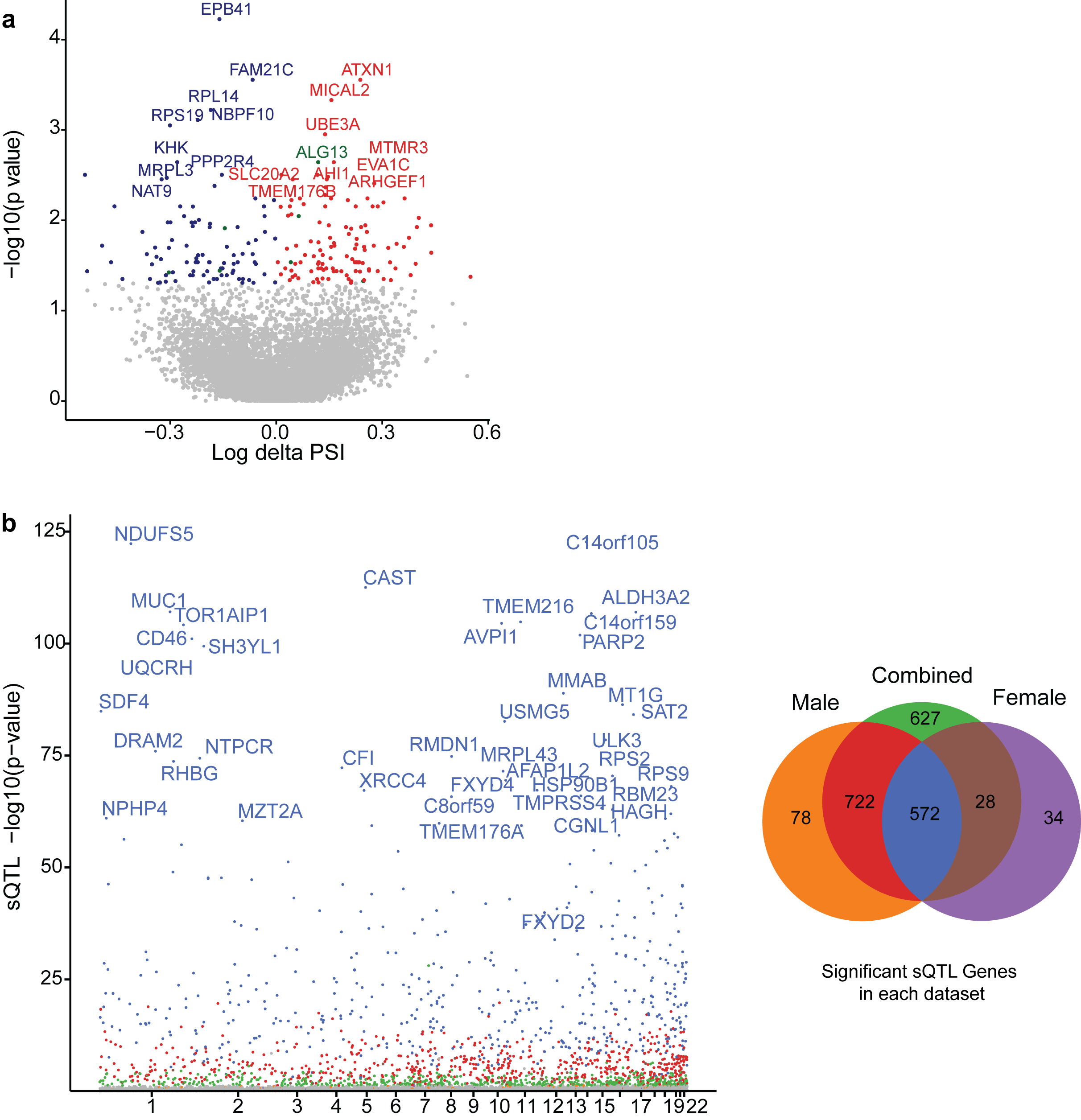

Supplementary Figure 9. Sex specific changes A. Delta PSI between male and female subjects demonstrating significant differences in splicing events by sex with 211 significant genes. x-axis shows the delta PSI between males and females, y axis the -log 10 adjusted p value. B. QTL analysis of male and females grouped separately in autologous chromosomes. X-axis is the location on the reference genome grouped by chromosomes. The SNPs are colored based on the venn diagram that shows the overlap of the genes. A small number of genes (78 and 34 in males and females respectively) are sex specific and masked in the entire cohort QTL.

Principal Investigators of the TRIDENT Consortium include Raymond Townsend, Gaia Coppock and Matthew Palmer (University of Pennsylvania), Manisha Singh (University of Arkansas Medical Center), Michael Ross (Albert Einstein College of Medicine), James Tumlin (Georgia Nephrology), Kirk Campbell (Mount Sinai School of Medicine), Amy Mottl (University of North Carolina), Christos Argyropoulos (University of New Mexico), Tamara Isakova (Northwestern University Feinberg School of Medicine), Salem Almaani (The Ohio State University), Rupali Avasare (Oregon Health and Science University), Richard Lafayette (Stanford University), Julia Scialla (University of Virginia), Randy Luciano (Yale University), Shweta Bansal (University of Texas Health Science Center at San Antonio), Frank Brosius (University of Arizona), Ankit Mehta (Baylor University), Oliver Lenz (University of Miami), Nelson Kopyt (Lehigh Valley Health Network), Piettro Canetta (Columbia University), Matthias Kretzler (University of Michigan), Jeffrey Schelling (Metro Health Medical Center), Alex Hofherr (AstraZeneca), Steven S Pullen (Boehringer Ingelheim Pharmaceuticals Inc), Andrew Thorley and David Kim (Genentech), John Liles (Gilead), Erding Hu (GlaxoSmithKline), Anil Karihaloo (Novo Nordisk Inc), and Kishor Devalaraja-Narashimha (Regeneron Pharmaceuticals Inc).
